# Cardiovascular and metabolic trajectories in men engaged in a multidimensional preventive care program: a prospective observational study

**DOI:** 10.64898/2026.11.12.26367899

**Authors:** Lisa Bedin, Maryne Lepoittevin, Paul Roujansky, Alaedine Benani, Rafik Gounane, Jerome Salomon, Emmanuel Sorberts, Gabriel Philippe Steg, Sylvain Bodard

## Abstract

**Objectives:** To describe within-individual changes in cardiometabolic, functional and neuroendocrine markers between two standardized evaluations in men engaged in a structured preventive care program, and to examine transitions across clinical risk categories.

**Design:** Prospective observational cohort study using routinely collected data; paired within-individual analyses.

**Setting:** Single preventive health center in Paris, France, offering a standardized multidisciplinary assessment with individualized lifestyle, supplementation and, when indicated, pharmacological recommendations.

**Participants:** 279 men aged 18 years or older with a first evaluation (CU1) between October 2022 and November 2025 and a second evaluation (CU2) by May 2026 (32.3% of 865 eligible men); median interval 13.3 months (IQR 12.0–15.6). No exclusion criteria were applied.

**Primary and secondary outcome measures:** Within-individual change in continuous markers (Hodges– Lehmann estimate, 95% CI; Benjamini–Hochberg adjustment over 36 tests) and transitions across fixed clinical thresholds (paired odds ratios).

**Results:** Median age was 51.2 years, BMI 24.5 kg/m² and SCORE2 2.9%. Visceral adipose tissue mass decreased (Δ −49 g, 95% CI −80.5 to −18.5; q=0.006), with small decreases in body mass and fat mass. Handgrip strength increased (right +2.35 kg, 1.70 to 3.00; left +1.70 kg, 1.05 to 2.30; q<0.001). HDL cholesterol increased (+3.5 mg/dL, 2.5 to 4.5; q<0.001), with decreases in the triglyceride/HDL and total/HDL ratios. Blood pressure, LDL cholesterol, fasting glucose (after adjustment), insulin, HOMA-IR, SCORE2 and salivary cortisol and DHEA did not change. The proportion of men below the grip strength reference value decreased from 60.1% to 47.6% (paired OR 0.34, 0.19 to 0.61) and with HDL cholesterol ≤46 mg/dL from 26.7% to 16.5% (OR 0.25, 0.12 to 0.54).

**Conclusions:** IIn this selected, health-engaged population of men who completed both evaluations, changes in a favorable direction were observed for handgrip strength, HDL cholesterol and visceral adiposity, whereas blood pressure, LDL cholesterol, glycemic markers and SCORE2 were stable. Without a comparison group, these changes cannot be distinguished from regression to the mean, test–retest effects or secular trends.

**Strengths and limitations of this study:**

- Standardized assessment protocol applied identically at both evaluations, combining clinical, laboratory, DEXA body composition, functional and salivary neuroendocrine measurements in the same individuals.
- Paired within-individual analyses of 36 markers in 279 men, reported as effect estimates with confidence intervals and a global false discovery rate adjustment, with sensitivity analyses by follow-up interval and calendar period and a comparison of returners with 586 eligible non-returners.
- Single-arm design without a comparison group: the observed changes cannot be distinguished from regression to the mean, test–retest effects, participation effects or secular trends.
- Highly selected population of urban, socioeconomically advantaged, health-engaged men, of whom 32% returned for a second evaluation, limiting generalizability to the general population and to high-risk groups.
- Standardized effect sizes were small for most markers, adherence to recommendations was not measured objectively, and the follow-up was short.

## Introduction

Cardiovascular disease accounts for nearly 20 million deaths annually, with a substantial proportion attributable to modifiable cardiometabolic risk factors including hypertension, dyslipidemia, hyperglycemia, and excess adiposity [1,2]. These risk factors frequently co-occur within individuals, interacting across vascular, metabolic, and inflammatory pathways in ways that amplify overall cardiovascular risk beyond the sum of individual contributions [3]. Recognizing this interdependence, contemporary preventive strategies increasingly emphasize integrated cardiometabolic assessment rather than the isolated management of single risk factors [4–6].

A multidimensional approach to cardiometabolic evaluation combines traditional clinical measures, such as blood pressure, lipid profiles, and glycemic markers, with complementary indicators that capture distinct but interrelated dimensions of risk. Visceral adiposity, for instance, is more closely linked to insulin resistance and systemic inflammation than overall body mass index alone [7,8]. Handgrip strength has emerged as an integrative marker of functional reserve independently associated with cardiovascular events and cause-specific mortality across extended follow-up periods [9–11]. Sleep disturbances and chronic psychosocial stress have also been increasingly linked to cardiometabolic dysregulation through neuroendocrine and behavioral mechanisms [12–14]. Composite risk algorithms such as SCORE2 further illustrate the value of integrating multiple determinants into absolute risk estimates to guide clinical decision-making [15,16]. Together, these domains form a broader cardiometabolic phenotype that may evolve dynamically over time and respond differently to preventive interventions.

However, most longitudinal evidence on cardiometabolic change derives from randomized trials of targeted interventions [4,17] or from population-based cohort studies evaluating isolated risk factors [18,19]. Relatively few studies have described how multiple cardiometabolic domains evolve simultaneously within individuals over short-term follow-up in structured, program-based preventive care settings. This gap is particularly relevant in the growing population of health-engaged adults who actively seek structured preventive pathways, a group that differs from both high-risk clinical cohorts and general population samples [20–22]. Understanding early multidimensional trajectories in these individuals may inform the design and calibration of personalized preventive strategies, yet such data remain scarce.

Two recent large observational studies have described cardiometabolic changes in adults exposed to structured prevention. In China, a digital-health multidomain lifestyle management programme followed 40,216 adults with home smart devices and wearables and reported greater reductions in systolic blood pressure, glucose and body fat percentage in those with high engagement [23]. In Japan, a retrospective analysis of consecutive annual health check-ups in adults with blood pressure, glucose or lipid abnormalities found one-year improvement in 29–57% of them, associated with weight loss and medication [24]. Neither study included research-grade body composition by DEXA, laboratory-measured lipid and glycaemic panels at both visits, or neuroendocrine measurements, and neither addressed adults voluntarily engaged in a programme-based preventive pathway in a European setting.

In this context, we analyzed data from a prospective cohort of adult men enrolled in an integrated preventive care program in France, combining standardized clinical, biological, imaging, and behavioral assessments with individualized preventive recommendations and digital follow-up. The primary objective was to describe within-individual changes in cardiometabolic risk markers between two consecutive standardized evaluations approximately one year apart. Secondary objectives were to describe these changes according to baseline risk profiles and to evaluate transitions across clinically relevant risk categories.

## Methods

### A. Study Design and Population

This prospective cohort study was based on clinical data collected during routine care at a preventive health center in Paris, France, and stored in the prospectively maintained cohort database whose design has been described in a cohort profile [25]. The study is reported according to the STROBE recommendations for cohort studies.

Men aged 18 years or older who attended the center for a first evaluation (Check-Up 1 [CU1]) between October 2022 and November 2025 and who had completed a second evaluation (Check-Up 2 [CU2]) at the time of data extraction (May 2026) were included. Participants enrolled either on their own initiative as self-paying individuals or through a programme offered by their employer; there was no recruitment for research purposes and no exclusion criteria were applied. The analysis was restricted to men as the first step of a sex-stratified analysis plan for the cohort, given sex differences in body composition, hormonal regulation and lipid trajectories [26] and the sex-specific SCORE2 algorithm [15];women will be analyzed separately (rationale detailed in the **Supplementary Methods**).

The preventive program (Zoī, Paris) is a structured multidisciplinary care pathway organized in four phases: a baseline assessment day (CU1) with standardized clinical, biological, imaging and behavioral evaluations; a follow-up medical consultation four weeks later, during which a physician reviews the results with the participant and delivers individualized preventive recommendations; a nurse-led follow-up call at three months; and an annual reassessment (CU2) using the same protocol as CU1. Between evaluations, participants had access to a smartphone application through which they could consult their results and recommendations, receive reminders and report whether recommendations had been followed; engagement metrics were not available for the present analysis. The program is described in detail in the **Supplementary Methods**.

Participants consented individually, before their first evaluation, that their pseudonymized health data could be used for research purposes and could object at any time, in accordance with French regulations for research on data collected during routine care (MR-004); no separate written consent was required for the present analysis. Data were processed in pseudonymized form and handled in compliance with applicable data protection requirements. The study was approved by the Ethics Committee of Sorbonne University (IRB00014283). Patient and public involvement: patients and the public were not involved in the design, conduct, reporting or dissemination plans of this research.

### B. Patient and Public Involvement statement

Patients and the public were not involved in setting the research question, the outcome measures, the design or the conduct of this study, which used data collected during routine care. Participants were informed individually of the research use of their data and could object

### C. Data Collection

#### a. Self-reported measures

##### i. Sociodemographic variables

Sociodemographic and lifestyle variables were collected at baseline using structured self-administered questionnaires: age, educational attainment, household income, professional status, alcohol consumption frequency, daily physical activity (yes/no), self-rated balanced diet (yes/no) and smoking status (never, former, current) (**Table 1**). Ethnicity and religion are not collected, in accordance with French law.

**Table 1:** Baseline characteristics of the study population at CU1 and interval between evaluations.

| Characteristic | Category / summary / value category | Value (N, %) |
|---|---|---|
| **Follow-up between CU1 and CU2 (months; n=276)** |  |  |
|  | **Mean (SD)** | 14.5 (4.2) |
|  | **Median [IQR]** | 13.3 [12.0–15.6] |
|  | **Range** | 3.1–37.6 |
| **Age at CU1 (years; n=276)** |  |  |
|  | **Mean (SD)** | 50.0 (10.8) |
|  | **Median [IQR]** | 51.2 [43.4–57.8] |
|  | **Range** | 18.9–84.7 |
| **Socio-demographics (n=279)** |  |  |
| **Education level** | Graduate or post-graduate | 226 (81.0 %) |
|  | Undergraduate | 26 (9.3 %) |
|  | Secondary | 19 (6.8 %) |
|  | Primary | 7 (2.5 %) |
|  | Not reported | 1 (0.4 %) |
| **Household income** | > €100 000 | 235 (84.2 %) |
|  | €35 000 – 100 000 | 25 (9.0 %) |
|  | ≤ €35 000 | 4 (1.4 %) |
|  | Prefer not to answer | 15 (5.4 %) |
| **Professional status** | Employee or head of a company | 225 (80.6 %) |
|  | Self-employed | 47 (16.8 %) |
|  | Unemployed | 7 (2.5 %) |
| **Lifestyle** |  |  |
| **Alcohol consumption (n=279)** | Daily | 34 (12.2 %) |
|  | Several times a week | 104 (37.3 %) |
|  | On weekends only | 63 (22.6 %) |
|  | Once to twice per month | 39 (14.0 %) |
|  | A few times a year | 14 (5.0 %) |
|  | Never | 25 (9.0 %) |
| **Daily physical activity (n=275)** | Yes | 193 (70.2 %) |
| **Balanced diet (n=279)** | Yes, most of the time | 203 (72.8 %) |
| **Smoking status (n=279)** | Never smoker | 186 (66.7 %) |
|  | Former smoker | 68 (24.4 %) |
|  | Current smoker | 23 (8.2 %) |
|  | Not recorded | 2 (0.7 %) |

*CU1, first check-up (baseline evaluation); CU2, second check-up (follow-up evaluation); SD, standard deviation; IQR, interquartile range (25th–75th percentile); n, number of participants with available data for the item; N, number of participants in the category. Percentages are calculated on the number of participants with available data for each item (denominator shown as n in the row header).*

##### ii. Sleep, Stress, and Perceived Health Measures

Risk of obstructive sleep apnea was screened using the STOP-BANG questionnaire (score 0–8, higher scores indicating greater risk) [27]; Polysomnography is not part of the initial assessment. **(Supplementary Methods**).

Daytime sleepiness was measured using the Epworth Sleepiness Scale (ESS, score 0–24) [28], with identical items at both visits. Perceived stress was assessed with a program questionnaire summarized as a score from 0 to 1 (**Supplementary Methods**); because this questionnaire was replaced by a different instrument between the two visits, the stress score is reported per visit and not compared between visits.

#### b. Clinical, biological, and functional assessments

Clinical and biological measurements were collected at CU1 and CU2. The complete list of longitudinally analyzed variables, their units and the number of participants with paired measurements is provided in **Table S1**.

##### i. Anthropometric measurements (Cardiometabolic anthropometry)

Body composition assessments included body weight, height, body mass index (BMI), waist circumference, total body fat mass, lean mass, visceral adipose tissue (VAT), and regional fat distribution measured by DEXA (GE, Lunar IDEXA,WI, USA). The android region is the DEXA-defined trunk segment between the pelvis and 20% of the distance to the neck; android fat/tissue mass is the fat mass of this region divided by its total soft-tissue mass, expressed as a percentage. Handgrip strength (right and left) was assessed using a calibrated digital dynamometer (KINVENT, K-Grip, Montpellier, France). Hemodynamic parameters included seated systolic and diastolic blood pressure and resting heart rate. These measurements were performed by a team of seven trained nurses. Detailed definitions of derived variables, categorical thresholds, and operational criteria for cardiometabolic conditions are provided in the **Supplementary Methods**.

##### ii. Biological measurements (Cardiometabolic Biochemical)

Participants attended the program in the morning in a fasting state, after a light dinner the evening before (typically rice, white meat and water) and nothing but water afterwards. The biomarker panel included lipid parameters (total cholesterol, HDL, calculated LDL and non-HDL cholesterol, triglycerides, the triglyceride/HDL ratio and the atherogenic index defined as total/HDL cholesterol), glycemic markers (fasting glucose, insulin [pmol/L], HbA1c [NGSP %], HOMA-IR), uric acid, and inflammatory markers (high-sensitivity C-reactive protein, homocysteine). Hs-CRP was added to the standard panel during the study period and was therefore not available at CU1 for all participants, which explains its smaller number of pairs.

##### iii. Stress biology (Sleep & Stress)

The day before the program, salivary cortisol was collected at awakening, 30 minutes post-awakening, and in the evening at home at 4°C; derived indices were the cortisol awakening response, the awakening-to-evening cortisol ratio and the evening cortisol/DHEA ratio (**Supplementary Methods**). The 10-year cardiovascular risk was estimated with SCORE2, which combines age, sex, smoking status, systolic blood pressure and non-HDL cholesterol, calibrated for low-risk countries including France. SCORE2 is validated for ages 40-69 years; it was recomputed at CU2 with the age and measurements of CU2, and the change is reported for the whole cohort and for men aged 40-69 years at both visits.

#### c. Preventive recommendations

Following the baseline preventive assessment (CU1), clinical and laboratory findings were reviewed by a physician and integrated into an individualized preventive care plan.

Recommendations were based on an integrated interpretation of clinical, DEXA, laboratory and patient-reported findings, and were explained and agreed with the participant in a shared decision-making approach. They included lifestyle advice (nutrition, physical activity, sleep, stress management), pharmacological treatment when clinically indicated (including statin therapy according to ESC/EAS LDL cholesterol targets), dietary supplements tailored to individual profiles, and follow-up monitoring; the indications for each supplement are given in the **Supplementary Methods**.

Implementation of the recommendations is recalled through the application notifications, and the three months nurse call. They are reviewed by the physician at CU2. However the data is only a subjective, declarative, followed/not followed status, documented for a minority of recommendations; changes in diet, physical activity and medication use between visits were not systematically measured. Adherence was therefore not analyzed.

### D. Variable Definition

Cardiometabolic conditions and interpretation categories were defined using established international clinical guidelines and standardized biomarker thresholds. Operational definitions, thresholds and calculation formulas are given in the **Supplementary Methods**.

### E. Outcomes

#### a. Primary Outcome

The primary outcome was the within-individual longitudinal change in continuous cardiometabolic variables between CU1 and CU2. These variables included clinical measurements, anthropometric and functional parameters, body composition measures derived from DEXA, blood and salivary biomarkers, SCORE2, and the STOP-BANG and Epworth scores.

#### b. Secondary Outcomes

Secondary outcomes examined complementary dimensions of longitudinal change. First, we described within-individual changes according to baseline categories of BMI, systolic blood pressure, glycemic status and LDL cholesterol. Second, we evaluated transitions between predefined clinical categories from CU1 to CU2.

All analyses relied on the same paired longitudinal framework as the primary analysis.

#### F. Statistical Analysis

Continuous variables were summarized as mean (SD) and median [interquartile range, IQR], and categorical variables as counts and percentages. Following current recommendations, results are presented as estimates with 95% confidence intervals (CI) and standardized effect sizes rather than in terms of statistical significance alone [29,30]; p-values are reported together with false discovery rate-adjusted q-values.

Before analysis, values outside pre-specified plausibility ranges (Supplementary Methods, section 5) were set to missing (14 values from 11 participants, e.g., visceral adipose tissue mass above 6,000 g or systolic blood pressure below 70 mmHg). Within-individual change between CU1 and CU2 was assessed in participants with non-missing values at both time points (complete-pair analysis; the number of pairs is given for each variable), for variables with at least 150 pairs. The paired difference was estimated by the Hodges-Lehmann estimator with its 95% CI derived from the Wilcoxon signed-rank distribution and the corresponding two-sided Wilcoxon signed-rank test; the mean difference (t-based 95% CI) and the paired Cohen’s d are also reported, and the relative change is the Hodges–Lehmann estimate divided by the CU1 median. Derived indices (triglyceride/HDL, total/HDL, non-HDL cholesterol, HOMA-IR) were recomputed from their constituents.

No sample size calculation was performed a priori; the sample corresponds to all eligible men in the database at extraction. A posteriori, with a median of 232 pairs per variable, a two-sided paired test at α=0.05 with 80% power detects a standardized difference d≈0.18 (d≈0.27 under a Bonferroni-type α for the 35 tests of **Table 2**). To assess selection into follow-up, men with a CU2 were compared with men whose CU1 took place at least 15 months before extraction but who had no CU2 (**Figure S1, Table S2**). Changes according to baseline category (**Table S3**) are descriptive and presented with confidence intervals only. Sensitivity analyses, detailed in the **Supplementary Methods**, separated the self-reported and objective STOP-BANG items (**Table S4**), repeated the **Table 2** analyses on pairs with a plausible interval of 6–24 months (**Table S5**), stratified blood and salivary biomarkers by period of the CU2 (**Table S6**), and examined the influence of the interval between evaluations (median 13.3 months) with annualized changes and the association between individual change and interval length (**Table S7**).

**Table 2:**
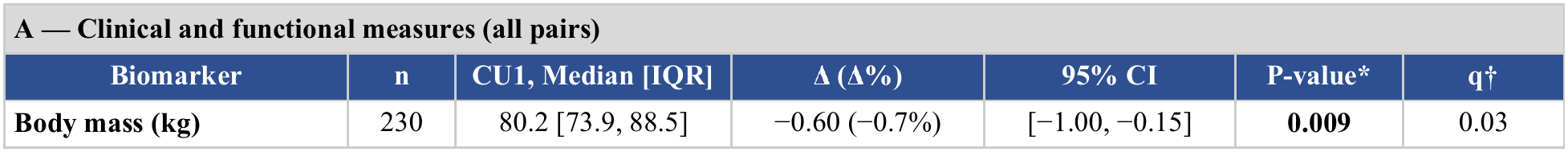
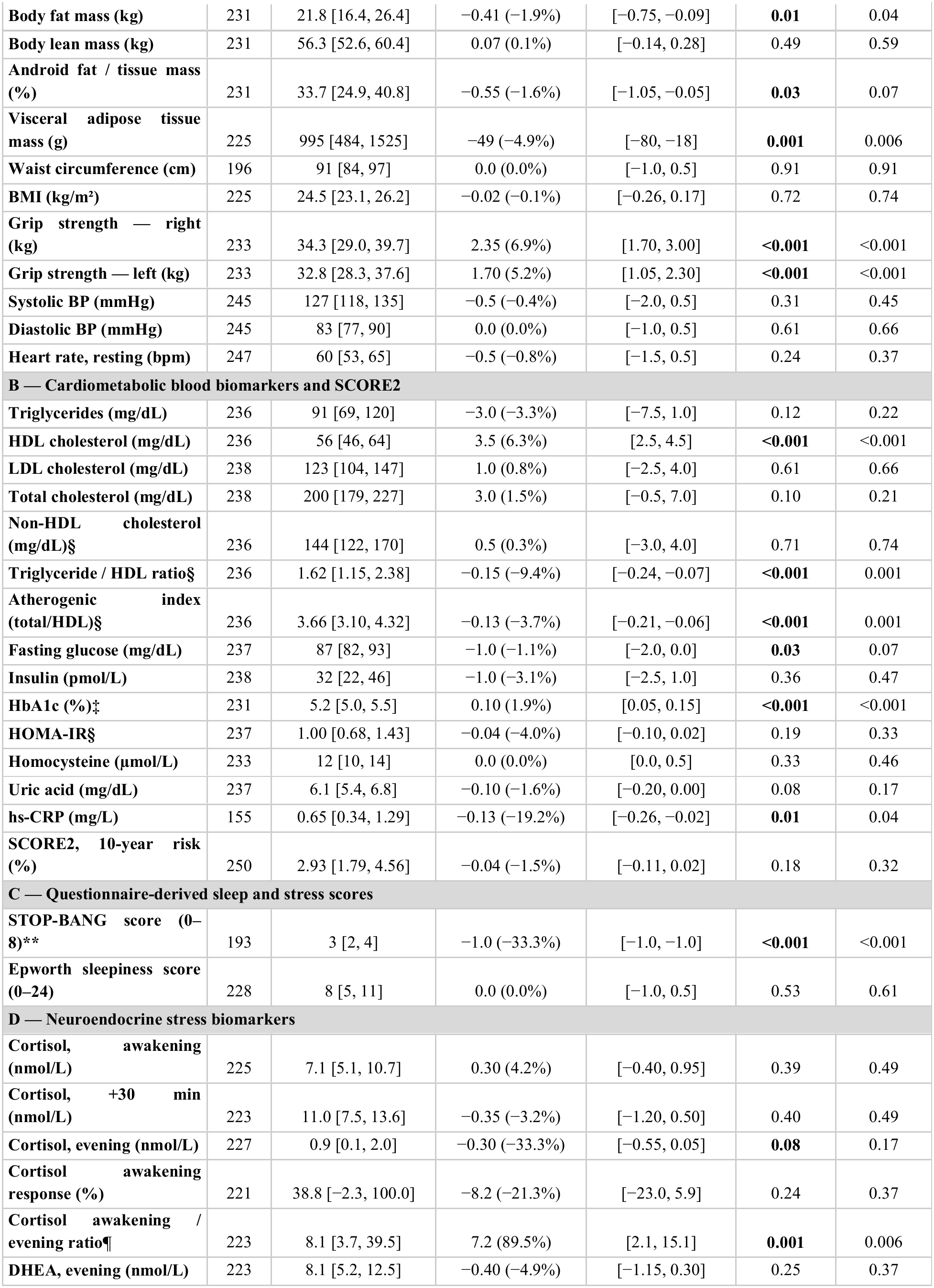
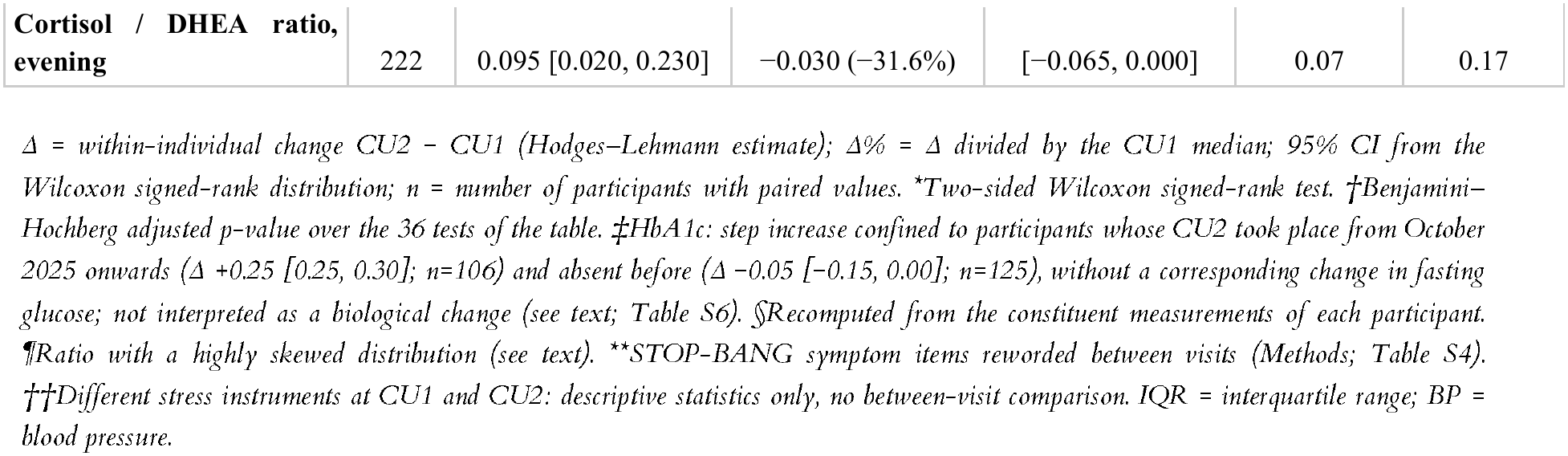
Within-individual changes in cardiometabolic, functional, questionnaire and neuroendocrine parameters between CU1 and CU2.

Transitions between clinical categories were analyzed with paired 2×2 tables: b denotes the number of participants moving from unfavorable at CU1 to favorable at CU2 and c the reverse; the paired odds ratio c/b (values below 1 indicating net improvement) is reported with a Wald 95% CI, the exact McNemar test and the net proportion improved (b−c)/n. These odds ratios are unadjusted by construction, each participant serving as his own control.

For multiple testing, p-values were adjusted with the Benjamini–Hochberg false discovery rate procedure applied globally over the 36 tests of Table 2 (families A to D together), and separately within Table 3 and within each supplementary table; the resulting q-values are shown next to the raw p-values. Analyses were conducted with R version (2025.05.1+513); the code and the analysis log are available from the corresponding author.

**Table 3.**
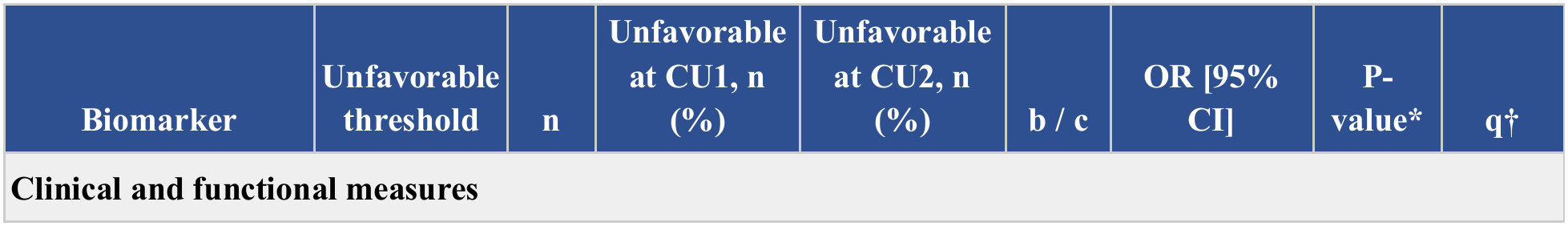
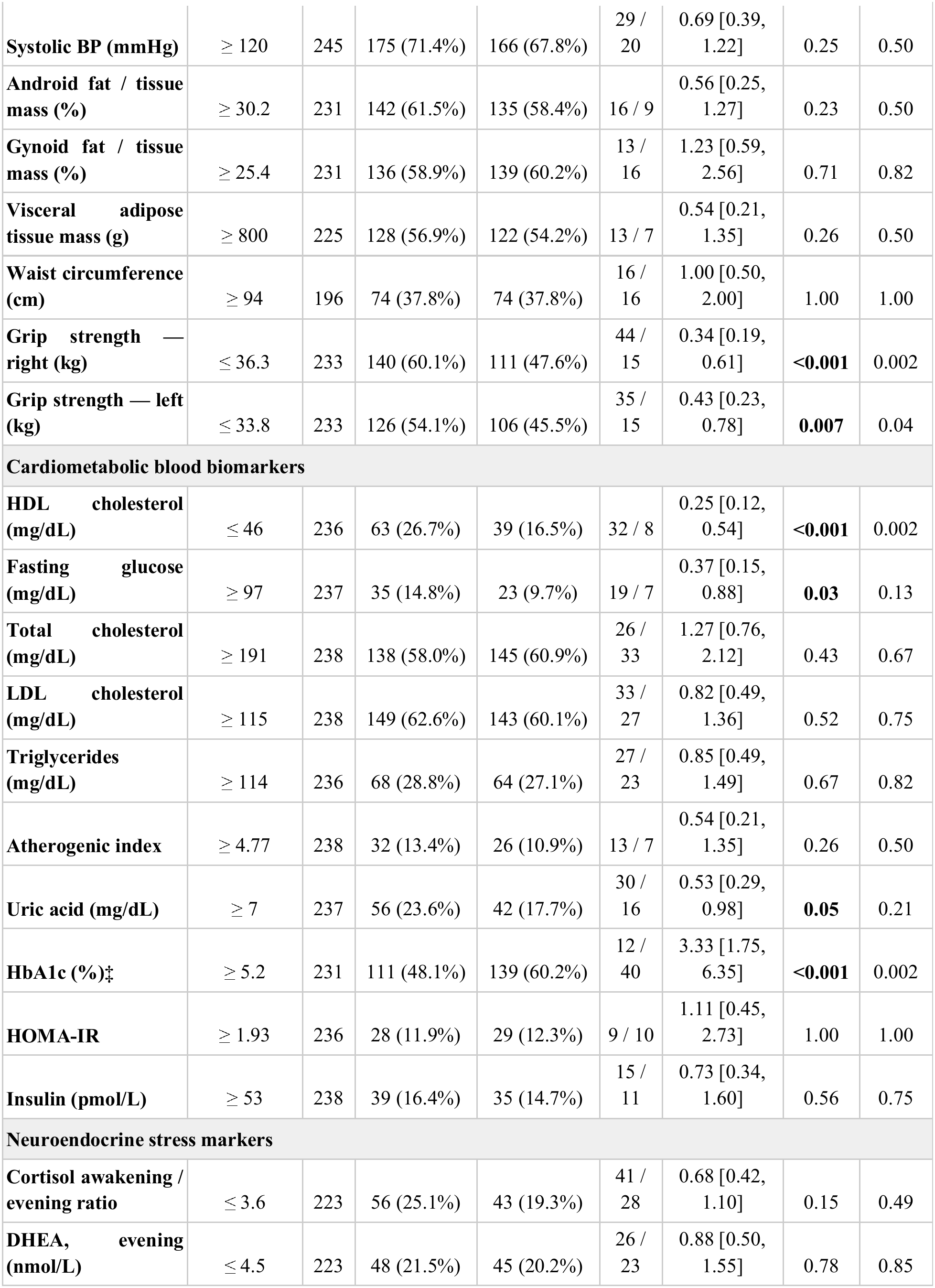
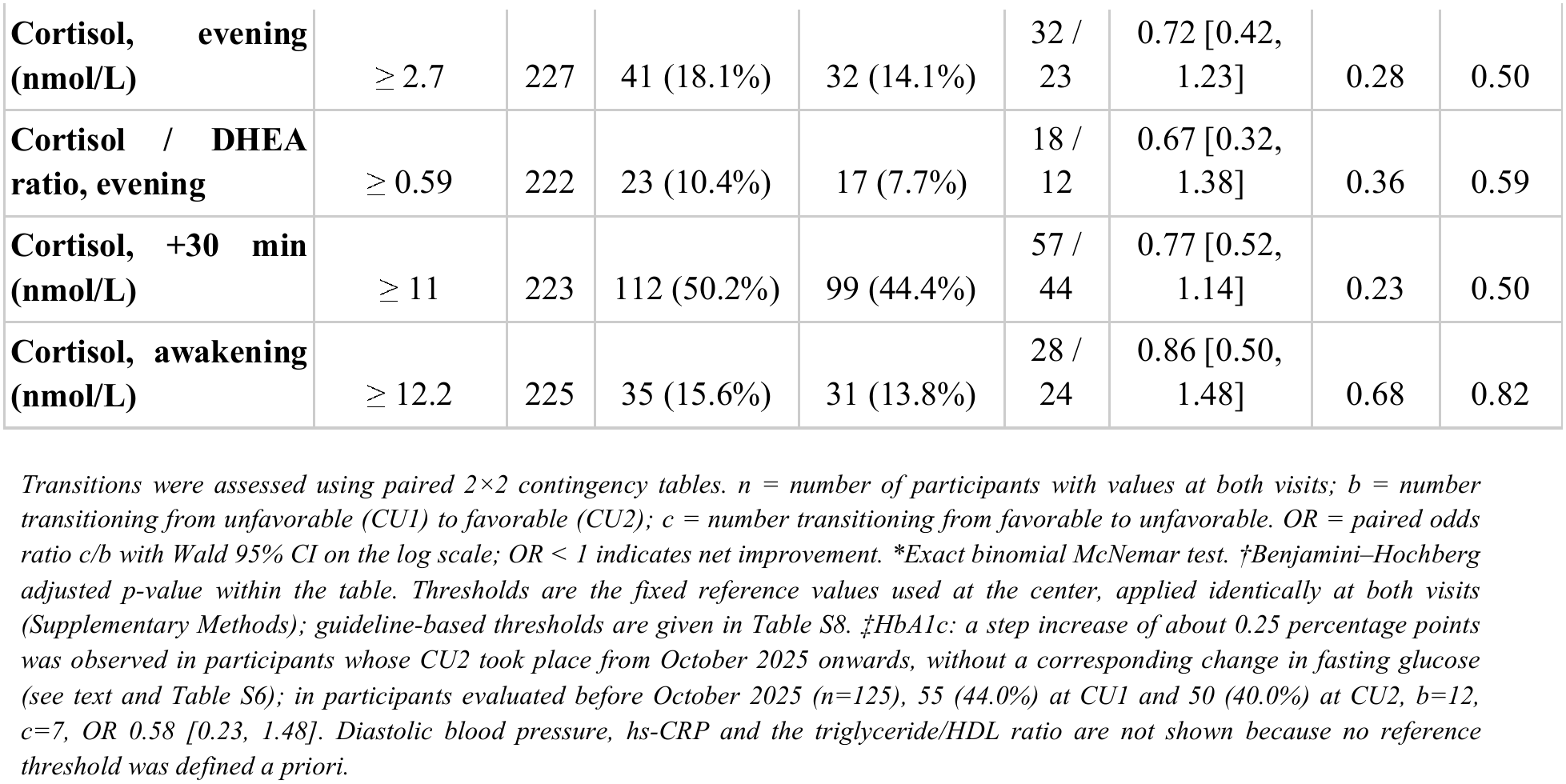
Transitions across clinical categories between CU1 and CU2 with paired odds ratios.

## Results

### A. Study Population and Follow-up

Of 2,871 adult men with a CU1 in the database, 865 had had their CU1 at least 15 months before data extraction and were therefore eligible for a CU2; 279 (32.3%) had completed a CU2 and constitute the analytical cohort (**Figure S1**). The interval between the two evaluations was 14.5 ± 4.2 months (median 13.3, IQR 12.0–15.6, range 3.1–37.6; n=276 with both dates recorded).The number of pairs differs between markers (155 to 250) because the assessment panel evolved over the study period and not every measurement was performed at both evaluations; it is given for each variable in the tables.Thirteen men had an interval shorter than 6 or longer than 24 months; all analyses of within-individual change were repeated without them and the estimates were unchanged ( **Supplementary Methods**). Mean age at CU1 was 50.0 ± 10.8 years (median 51.2, range 18.9–84.7). Sociodemographic questionnaires were completed by all participants: 226 (81.0%) reported graduate-level education, 235 (84.2%) an annual household income above €100 000, 193 of 275 (70.2%) daily physical activity and 203 (72.8%) a balanced diet most of the time; 23 (8.2%) were current smokers and 68 (24.4%) former smokers. Compared with eligible men who had not returned for a CU2 (n=586), participants did not differ meaningfully in age, BMI, blood pressure, lipids, glycemic markers, visceral adipose tissue, grip strength or SCORE2 at CU1 (all absolute standardized mean differences ≤0.14; **Table S2**). Baseline characteristics are presented in **Table 1**.

### B. Changes in Cardiometabolic Parameters between CU1 and CU2

At baseline, the cohort presented a low-to-moderate cardiometabolic risk profile. Median BMI was 24.5 kg/m² (IQR 23.1–26.2). Among the 225 participants with paired data, 79 (35.1%) were overweight and 15 (6.7%) obese. Median systolic blood pressure was 127 mmHg (IQR 118–135); 61 of 245 (24.9%) had high-normal values (130–139 mmHg) and 34 (13.9%) values ≥140 mmHg. Median fasting glucose was 87 mg/dL (IQR 82–93) and 30 of 237 participants (12.7%) had fasting glucose ≥100 mg/dL or HbA1c ≥5.7%. Median LDL cholesterol was 123 mg/dL (IQR 104–147) and median 10-year SCORE2 risk 2.9% (IQR 1.8–4.6). Baseline values, numbers of pairs and within-individual changes for all markers are presented in Table 2.

#### a. Clinical and functional measures

Among body composition markers, visceral adipose tissue mass decreased (Δ −49 g, 95% CI −80.5 to −18.5; −4.9% of the baseline median; q=0.006), with small decreases in body mass (Δ −0.60 kg, −1.00 to −0.15; q=0.03) and body fat mass (Δ −0.41 kg, −0.75 to −0.09; q=0.04). The android fat/tissue mass ratio (Δ −0.55 percentage points, −1.05 to −0.05; q=0.07), lean mass, BMI and waist circumference did not change. Handgrip strength increased bilaterally (right: Δ +2.35 kg, 1.70 to 3.00; left: Δ +1.70 kg, 1.05 to 2.30; both q<0.001). Systolic and diastolic blood pressure and resting heart rate did not change (**Table 2, A**).

In descriptive subgroup analyses (**Table S3, A**), BMI decreased in participants with obesity at baseline (BMI ≥30 kg/m²; n=15; Δ −1.75 kg/m², 95% CI −2.69 to −0.69), but not in those with overweight (n=79; Δ −0.14, −0.55 to 0.23) or normal weight (n=131; Δ +0.16, −0.03 to 0.40). Systolic blood pressure decreased in participants with high-normal values at baseline (130–139 mmHg; n=61; Δ −3.5 mmHg, −5.5 to −0.5) and in those with values ≥140 mmHg (n=34; Δ −4.5 mmHg, −9.0 to −1.0), and tended to increase in those below 130 mmHg (n=150; Δ +1.0, −0.00 to 2.5), a pattern compatible with regression to the mean.

#### b. Cardiometabolic blood biomarker

HDL cholesterol increased (Δ +3.5 mg/dL, 95% CI 2.5 to 4.5; n=236; q<0.001), with decreases in the triglyceride/HDL ratio (Δ −0.15, −0.24 to −0.07; q=0.001) and the total/HDL cholesterol ratio (Δ −0.13, −0.21 to −0.06; q=0.001). Triglycerides (Δ −3.0 mg/dL, −7.5 to 1.0; q=0.22), total, LDL and non-HDL cholesterol did not change. Fasting glucose showed a small decrease that did not survive adjustment for multiple testing (Δ −1.0 mg/dL, −2.0 to 0.0; q=0.07), and insulin and HOMA-IR (Δ −0.04, −0.10 to 0.02; q=0.33) did not change. HbA1c increased by 0.10 percentage points (0.05 to 0.15; q<0.001); this change was confined to participants whose CU2 took place from October 2025 onwards (Δ +0.25, 0.25 to 0.30; n=106) and was absent in those evaluated earlier (Δ −0.05, −0.15 to 0.00; n=125), with no corresponding difference in fasting glucose, and is therefore not interpreted as a biological change (**Table S6**). hs-CRP decreased (Δ −0.13 mg/L, −0.26 to −0.02; n=155; q=0.04), whereas uric acid and homocysteine were stable. SCORE2 did not change (Δ −0.04 percentage points, −0.11 to 0.02; q=0.32); in men aged 40–69 years at both visits (n=198) the estimate was −0.08 (−0.16 to 0.00; p=0.049, d=−0.12, not adjusted for multiple testing), a difference of less than 0.1 percentage point that we do not regard as meaningful (**Table 2, B**). Estimates on pairs with a plausible interval were similar (**Table S5**); estimates by period of the CU2 are given in **Table S6**.

In descriptive subgroup analyses (**Table S3, B**), fasting glucose changed by −1.0 mg/dL (95% CI −2.0 to 0.0) in normoglycemic participants (n=207) and by −2.5 mg/dL (−6.5 to 0.5) in the 30 participants with fasting glucose ≥100 mg/dL or HbA1c ≥5.7% at baseline. LDL cholesterol increased in participants with baseline LDL <116 mg/dL (n=95; Δ +6.5 mg/dL, 2.0 to 11.0), did not change in the 116–189 mg/dL category (n=133; Δ −2.0, −7.5 to 2.5) and decreased in the 10 participants with LDL ≥190 mg/dL (Δ −38.5 mg/dL, −78.0 to −4.0); this last estimate rests on 10 observations, none of whom had a statin prescription recorded at CU1; a statin may however have been initiated between the two evaluations following the medical review, which is not captured in the database, and the estimate is reported for description only.

#### c. Sleep, stress, and neuroendocrine markers

The STOP-BANG score decreased by a median of one point (Hodges–Lehmann Δ −1.0; mean Δ −0.59, 95% CI −0.73 to −0.45; n=193; d=−0.59; q<0.001). This decrease was entirely carried by the three self-reported symptom items (S+T+O: Δ −1.0; n=193), whose wording was revised between visits, whereas the objective items (B+A+N+G+P) did not change (Δ 0.0, 95% CI −1.0 to 0.0; d=−0.09; n=193) (Table S4). The Epworth sleepiness score did not change (Δ 0.0, 95% CI −1.0 to 0.5; n=228). Among neuroendocrine markers, awakening, post-awakening and evening cortisol, the cortisol awakening response, evening DHEA and the cortisol/DHEA ratio did not change. The awakening-to-evening cortisol ratio increased (Δ +7.2, 95% CI 2.1 to 15.1; q=0.006); this ratio has a highly skewed distribution (baseline IQR 3.7–40) driven by very low evening values, and its change should be regarded as hypothesis-generating (Table 2, C–D).

### C. Transitions Across Clinical Risk Categories

To complement the continuous analyses, we examined transitions between favorable and unfavorable categories defined by fixed thresholds, using paired 2×2 tables (**Table 3; Table S8**). Paired individual values for six markers are shown in Figure 1.

**Figure 1.**
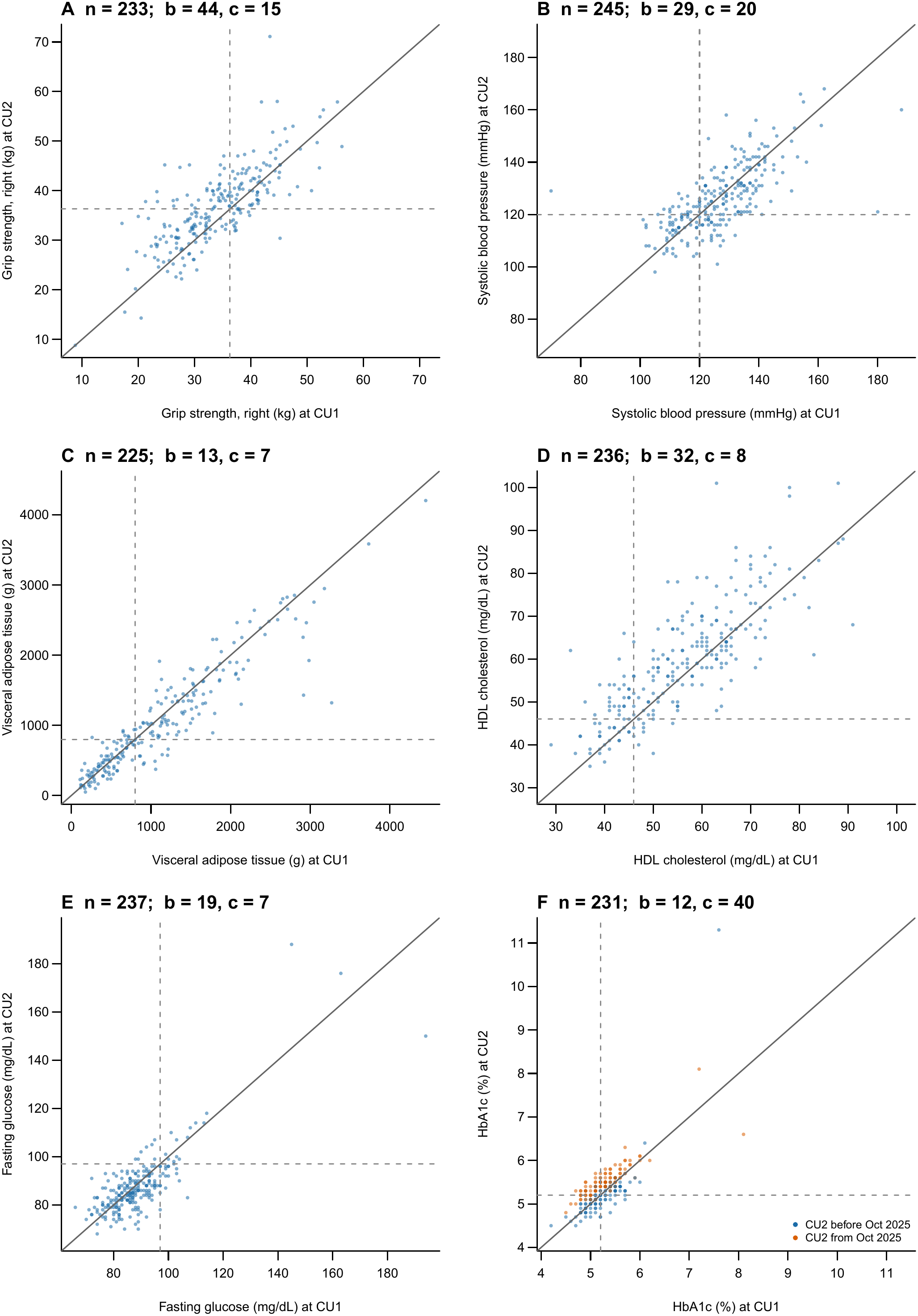
Paired individual values at CU1 (x□axis) and CU2 (y□axis) for six markers.

For right-hand grip strength, the proportion of participants below the reference value (≤36.3 kg) decreased from 60.1% at CU1 to 47.6% at CU2 (44 participants improved versus 15 who worsened; paired OR 0.34, 95% CI 0.19 to 0.61; q=0.002), and for the left hand from 54.1% to 45.5% (35 versus 15; OR 0.43, 0.23 to 0.78; q=0.04). The proportions with systolic blood pressure ≥120 mmHg (71.4% to 67.8%; OR 0.69, 0.39 to 1.22), visceral adipose tissue mass ≥800 g (56.9% to 54.1%; OR 0.54, 0.21 to 1.35), android or gynoid fat ratio above threshold, or waist circumference ≥94 cm did not change beyond chance.

Among blood biomarkers, the proportion with HDL cholesterol ≤46 mg/dL decreased from 26.7% to 16.5% (32 improved versus 8 worsened; OR 0.25, 95% CI 0.12 to 0.54; q=0.002). The proportion with fasting glucose ≥97 mg/dL decreased from 14.8% to 9.7% (19 versus 7; OR 0.37, 0.15 to 0.88; q=0.13) and the proportion with uric acid ≥7.0 mg/dL from 23.6% to 17.7% (30 versus 16; OR 0.53, 0.29 to 0.98; q=0.21). No net transition was observed for total or LDL cholesterol, triglycerides, the atherogenic index, HOMA-IR or insulin. The proportion with HbA1c ≥5.2% increased from 48.1% to 60.2% (12 versus 40; OR 3.33, 1.75 to 6.35); this transition reflects the period discontinuity described above and was not observed in participants whose CU2 preceded October 2025 (44.0% to 40.0%; 12 versus 7; OR 0.58, 0.23 to 1.48; n=125).

Among neuroendocrine markers, no transition differed from chance (Table 3).

## Discussion

This study describes within-individual changes in cardiometabolic, functional, questionnaire-based and neuroendocrine markers between two standardized evaluations, a median of 13 months apart, in 279 men engaged in a structured preventive care program. Handgrip strength and HDL cholesterol increased, the triglyceride/HDL and total/HDL cholesterol ratios, visceral adipose tissue mass, body mass, fat mass and hs-CRP decreased. Blood pressure, LDL cholesterol, fasting glucose (after adjustment for multiple testing), insulin, HOMA-IR, SCORE2 and salivary cortisol and DHEA did not change. An increase in HbA1c was observed but is confined to a specific calendar period and is not interpreted as a biological change. Standardized effect sizes were small for all markers (|d| ≤ 0.29) except grip strength and HDL cholesterol (d 0.43–0.48); the changes described here are therefore modest at the individual level, whatever their direction.These changes, observed in men who completed both evaluations, are in the direction expected from the recommendations delivered; a comparison group will be needed to determine how much of them is attributable to the program. Across the four domains examined, the direction of change was consistently favorable or neutral, with no marker moving in an unfavorable direction.

### Body composition and grip strength

Visceral adipose tissue mass decreased by 49 g (about 5% of the baseline median) while BMI and waist circumference were stable and body mass and fat mass decreased by about half a kilogram. Visceral adiposity is considered a more relevant metabolic correlate of cardiovascular risk than BMI alone [7]; the changes observed here are small at the individual level (Cohen’s d ≤ 0.25) and their clinical relevance over one year is uncertain. Handgrip strength increased bilaterally (right +2.35 kg, left +1.70 kg, about 5–7% of the baseline median), and the proportion of men below the reference value for the right hand decreased from 60% to 48%. Grip strength is an integrative marker of functional reserve and an independent predictor of cardiovascular events and mortality [9–11], and it declines by about 0.5–0.7 kg per year in men over 50 in population-based studies [31]. However, a second measurement with the same device after one year is susceptible to familiarization and motivation effects: test–retest reliability of grip dynamometry is high but not perfect [32], and the increase observed here (about 2 kg) is of the order of the minimal clinically important differences reported for grip strength (about 5–6 kg in most studies)[33]. The increase was also smaller in men with longer intervals between evaluations (**Table S7**). The observed increase can therefore not be interpreted as a functional improvement attributable to the program..

### Lipid profile and estimated cardiovascular risk

HDL cholesterol increased by 3.5 mg/dL (6% of the baseline median), with corresponding decreases in the triglyceride/HDL ratio and the atherogenic index, whereas total, LDL and non-HDL cholesterol and triglycerides did not change. SCORE2 did not change either (Δ −0.04 percentage points, 95% CI −0.11 to 0.02), with a similar estimate (−0.08, −0.16 to 0.00) in the 40–69-year age range for which the algorithm is validated. Because SCORE2 increases mechanically with age, a stable estimate over 13 months indicates that small changes in its components (systolic blood pressure, non-HDL cholesterol, smoking status) offset the effect of ageing over the interval; it does not indicate that risk was reduced. The stability of LDL cholesterol at the cohort level hides opposite trajectories by baseline category (**Table S3**): LDL increased in men with baseline values below 116 mg/dL, did not change in the 116–189 mg/dL category and decreased in the 10 men with values ≥190 mg/dL. This pattern is the expected signature of regression to the mean in extreme baseline categories [27]; none of the 10 men had a statin prescription recorded at CU1, and medication use between visits was not systematically recorded, so that a treatment effect can neither be invoked nor excluded.

### Glycemic and inflammatory markers

Fasting glucose decreased by 1 mg/dL, a change that did not survive adjustment for multiple testing (q=0.07), and the proportion of men with fasting glucose ≥97 mg/dL decreased from 15% to 10% (q=0.13); insulin and HOMA-IR did not change. HbA1c increased by 0.10 percentage points, but this increase is entirely due to a step of about 0.25 points in men whose CU2 took place from October 2025 onwards, with no change earlier and no corresponding change in fasting glucose (Table S6). A homogeneous shift of this size confined to a calendar period is not compatible with a biological change in glycemic control; HbA1c results are therefore reported but not interpreted. hs-CRP decreased by 0.13 mg/L (about 19% of a low baseline median of 0.65 mg/L); given the skewed distribution of this marker and the absence of a corresponding change in body composition of comparable magnitude, this small change is reported without interpretation.

### Observed changes in the context of expected age-related trajectories

Several markers that are expected to deteriorate with age, grip strength [31];SCORE2, fasting glucose [37]

### Sleep, stress and neuroendocrine markers

The STOP-BANG score decreased by a median of one point. The Epworth score did not change. The perceived-stress questionnaire was replaced by a different instrument between the visits and its scores were not compared. Salivary cortisol and DHEA measurements were stable; the increase in the awakening/evening cortisol ratio is driven by very low evening values in the denominator of a highly skewed ratio, no neuroendocrine transition differed from chance (**Table 3**), and this isolated finding is regarded as hypothesis-generating only.

### Categorical transitions

After adjustment for multiple testing, only two transitions differed from chance: HDL cholesterol (proportion with values ≤46 mg/dL from 27% to 17%) and right-hand grip strength (proportion below the reference value from 60% to 48%), with left-hand grip strength at the margin. Transitions for systolic blood pressure, visceral adipose tissue, fasting glucose and uric acid were in a favorable direction but compatible with chance, and no transition was observed for LDL cholesterol, HOMA-IR or insulin. The apparent unfavorable transition for HbA1c reflects the period discontinuity discussed above and disappears in men evaluated before October 2025. These transitions are unadjusted paired comparisons and share the limitations of the continuous analyses.

### Comparison with other program-based cohorts

Two recent large observational studies have described cardiometabolic changes in adults exposed to structured prevention. In the digital-health cohort of Xiao et al. [23], 40,216 Chinese adults followed with connected devices showed greater reductions in systolic blood pressure, glucose and body fat percentage with higher engagement; in the Japanese check-up cohort of Kameda et al. [24], 29–57% of adults with blood pressure, glucose or lipid abnormalities had improved one year later, in association with weight loss and medication. Our cohort is smaller and younger, at lower baseline risk, and evaluated with research-grade measurements (DEXA, laboratory panels at both visits, salivary neuroendocrine markers) rather than home devices or routine check-up data. The direction and magnitude of the changes we observe in blood pressure and lipids are compatible with these reports, with the same interpretive limits, since none of the three studies includes a comparison group.

### Digital tools in preventive care

Participants had access to a smartphone application through which they could consult their results and recommendations and report whether recommendations had been followed. Engagement metrics were not available for the present analysis, so that the relation between digital engagement and the observed changes could not be examined. Digital tools (applications, wearables, step-count monitoring, remote coaching and automated feedback) are increasingly proposed as scalable components of cardiometabolic prevention, and the cohort of Xiao et al. suggests that the level of engagement matters [23]. Whether objectively measured engagement and activity patterns contribute to trajectories such as those observed here is a priority for future analyses of the cohort.

### Restriction to male participants

The analysis was restricted to men as the first step of a sex-stratified analysis plan for the cohort, not as a convenience sample. Sex differences in body composition, hormonal regulation and lipid trajectories across the life course are well documented [26,38], SCORE2 is sex-specific [16] and the analysis of women requires accounting for menopausal status and for cyclic variation of some inflammatory and neuroendocrine markers. Stratifying a mixed cohort would equally have avoided confounding by sex; analyzing the two sexes in separate reports was chosen because the outcome definitions and the interpretation differ. The analysis of women is in preparation.

### Several strengths of this study should be noted

First, data were collected within a structured preventive care center using a standardized assessment protocol applied identically at both evaluations. Second, the study draws on a prospectively maintained clinical database whose design has been reported in a cohort profile [39], combining clinical, biological, body composition (DEXA), functional, questionnaire and neuroendocrine data on the same individuals, a combination that neither of the two recent large program-based cohorts provides [23,24]. Third, the real-world design captures cardiometabolic dynamics as they occur in routine preventive practice. Fourth, the analysis applies pre-specified plausibility rules, a single consistent estimator with its confidence interval and test, a global correction for multiple testing, and several sensitivity analyses, and reports the number of pairs for every variable. Finally, the ongoing nature of the cohort will allow the evaluation of longer-term trajectories.

### Several limitations should be acknowledged

First, the absence of a comparison group precludes attributing the observed changes to the program rather than to natural evolution, regression to the mean [36], test–retest effects, secular trends or the behaviour change induced by observation itself (Hawthorne effect) [40]; participants knew they would be re-evaluated, health check-ups by themselves are associated with subsequent changes in risk factors although not with reduced morbidity or mortality in randomized trials [35], and the magnitude of these effects cannot be quantified in a single-arm design. Second, the study population, men attending a preventive center in Paris, 81% with graduate education, 84% with a household income above €100 000, 8% current smokers compared with about a quarter of French men [41], and a median BMI of 24.5 kg/m², represents neither the general French population nor the high-risk populations usually targeted by prevention programs [20–22], which limits generalizability. Third, only 32% of eligible men returned for a second evaluation. Their baseline characteristics were close to those of non-returners (all standardized mean differences ≤0.14; **Table S2**), so that selection on baseline risk appears limited, but selection on motivation or on the trajectory itself cannot be excluded and would bias changes in a favorable direction. Fourth, adherence to recommendations was captured only as a self-reported followed/not followed status, documented for a minority of recommendations, and changes in diet, physical activity and medication use between evaluations were not measured; engagement metrics of the digital platform were not available for this analysis. Associations between recommendations and biomarker changes could therefore not be analyzed, and the behavioural pathways through which the observed changes may have occurred cannot be assessed. Fifth, STOP-BANG and stress questionnaires were revised between the visits, and all questionnaire scores are self-reported and susceptible to reporting bias. Sixth, the interval between evaluations varied (3 to 38 months, median 13.3); changes were unrelated to the interval for almost all markers and the results were unchanged when restricted to intervals of 6–24 months (Tables S5 and S7), but the annualized changes should be read with this heterogeneity in mind. Finally, the follow-up is short and the analysis involves 36 paired comparisons; despite the false discovery rate adjustment, the smaller changes should be regarded as descriptive, and none of the observed changes can be related to clinical events.

## Conclusion

In this prospective cohort of 279 men engaged in a structured preventive care program, within-individual changes between two standardized evaluations a median of 13 months apart were in a favorable direction for handgrip strength, HDL cholesterol and its ratios, visceral adipose tissue, body mass and fat mass, whereas blood pressure, LDL cholesterol, glycemic markers, SCORE2 and salivary neuroendocrine markers were stable. These changes are small at the individual level, were observed in a selected, health-engaged population without a comparison group, and cannot be attributed to the program rather than to regression to the mean, test–retest effects, participation effects or secular trends. The study illustrates the multidimensional cardiometabolic trajectories that can be described within a program-based preventive setting and the data-quality requirements of such descriptions in routine care. Establishing whether programs of this type modify cardiometabolic trajectories will require a comparison group, objective measures of adherence and digital engagement, longer follow-up and the inclusion of women.

## Data Availability

All data produced in the present study are available upon reasonable request to the authors

## Acknowledgements

The authors express their gratitude to the entire Zoī team, including physicians, nurses, clinical staff, data engineers, and administrative personnel, for their dedication to the preventive care program and their contribution to the collection and management of the data used in this study.

Use of Artificial Intelligence. During the preparation of this manuscript, the authors used Claude (Opus 5, Anthropic) to assist with manuscript drafting, editing, and formatting. All AI-generated content was reviewed, verified, and edited by the authors, who take full responsibility for the content of the published article.

## Ethics approval

The study was approved by the Ethics Committee of the Sorbonne Université (IRB accreditation no.IRB00014283)

## Consent to participate

The study involved only pseudonymized data. All participants were informed generally and individually prior to inclusion, with the possibility to object, in accordance with national regulations.

## Consent to publish

No individual-identifiable data or images are included in this manuscript.

## References

1 The Global Burden of Cardiovascular Diseases and Risks: A Compass for Global Action | JACC. https://www.jacc.org/doi/10.1016/j.jacc.2020.11.021 (accessed 20 February 2026)

2 Vos T, Lim SS, Abbafati C, et al. Global burden of 369 diseases and injuries in 204 countries and territories, 1990–2019: a systematic analysis for the Global Burden of Disease Study 2019. The Lancet. 2020;396:1204–22. doi: 10.1016/S0140-6736(20)30925-9

3 Alberti KGMM, Eckel RH, Grundy SM, et al. Harmonizing the metabolic syndrome: a joint interim statement of the International Diabetes Federation Task Force on Epidemiology and Prevention; National Heart, Lung, and Blood Institute; American Heart Association; World Heart Federation; International Atherosclerosis Society; and International Association for the Study of Obesity. Circulation. 2009;120:1640–5. doi: 10.1161/CIRCULATIONAHA.109.192644

4 William C Knowler. Reduction in the Incidence of Type 2 Diabetes with Lifestyle Intervention or Metformin. New England Journal of Medicine. 2002;346:393–403. doi: 10.1056/NEJMoa012512

5 Primary Prevention of Cardiovascular Disease with a Mediterranean Diet. New England Journal of Medicine. 2014;370:886–886. doi: 10.1056/NEJMx140004

6 Tsao CW, Aday AW, Almarzooq ZI, et al. Heart Disease and Stroke Statistics—2023 Update: A Report From the American Heart Association. Circulation. 2023;147:e93–621. doi: 10.1161/CIR.0000000000001123

7 Body Fat Distribution and Risk of Cardiovascular Disease | Circulation. https://www.ahajournals.org/doi/10.1161/CIRCULATIONAHA.111.067264?url_ver=Z39.88-2003&rfr_id=ori:rid:crossref.org&rfr_dat=cr_pub%20%200pubmed (accessed 13 March 2026)

8 Wormser D, Wood AM, Angelantonio ED, et al. Metabolic mediators of body-mass index and cardiovascular risk. The Lancet. 2014;383:2042–3. doi: 10.1016/S0140-6736(14)60991-0

9 Celis-Morales CA, Welsh P, Lyall DM, et al. Associations of grip strength with cardiovascular, respiratory, and cancer outcomes and all cause mortality: prospective cohort study of half a million UK Biobank participants. BMJ. 2018;361:k1651. doi: 10.1136/bmj.k1651

10 Sayer AA, Kirkwood TBL. Grip strength and mortality: a biomarker of ageing? The Lancet. 2015;386:226–7. doi: 10.1016/S0140-6736(14)62349-7

11 Sasaki H, Kasagi F, Yamada M, et al. Grip Strength Predicts Cause-Specific Mortality in Middle-Aged and Elderly Persons. The American Journal of Medicine. 2007;120:337–42. doi: 10.1016/j.amjmed.2006.04.018

12 Sleep Duration and Quality: Impact on Lifestyle Behaviors and Cardiometabolic Health: A Scientific Statement From the American Heart Association. Circulation.

13 Cappuccio FP, D’Elia L, Strazzullo P, et al. Sleep Duration and All-Cause Mortality: A Systematic Review and Meta-Analysis of Prospective Studies. Sleep. 2010;33:585–92. doi: 10.1093/sleep/33.5.585

14 Chandola T, Britton A, Brunner E, et al. Work stress and coronary heart disease: what are the mechanisms? Eur Heart J. 2008;29:640–8. doi: 10.1093/eurheartj/ehm584

15 SCORE2 working group and ESC Cardiovascular risk collaboration, Hageman S, Pennells L, et al. SCORE2 risk prediction algorithms: new models to estimate 10-year risk of cardiovascular disease in Europe. European Heart Journal. 2021;42:2439–54. doi: 10.1093/eurheartj/ehab309

16 2021 ESC Guidelines on cardiovascular disease prevention in clinical practice | European Heart Journal | Oxford Academic. https://academic.oup.com/eurheartj/article/42/34/3227/6358713 (accessed 20 February 2026)

17 The Look AHEAD Research Group. Cardiovascular Effects of Intensive Lifestyle Intervention in Type 2 Diabetes. New England Journal of Medicine. 2013;369:145–54. doi: 10.1056/NEJMoa1212914

18 Yusuf S, Hawken S, Ôunpuu S, et al. Effect of potentially modifiable risk factors associated with myocardial infarction in 52 countries (the INTERHEART study): case-control study. The Lancet. 2004;364:937–52. doi: 10.1016/S0140-6736(04)17018-9

19 Collaboration TERF. Diabetes mellitus, fasting blood glucose concentration, and risk of vascular disease: a collaborative meta-analysis of 102 prospective studies. The Lancet. 2010;375:2215–22. doi: 10.1016/S0140-6736(10)60484-9

20 Havranek EP, Mujahid MS, Barr DA, et al. Social Determinants of Risk and Outcomes for Cardiovascular Disease. Circulation. 2015;132:873–98. doi: 10.1161/CIR.0000000000000228

21 Schultz WM, Kelli HM, Lisko JC, et al. Socioeconomic Status and Cardiovascular Outcomes. Circulation. 2018;137:2166–78. doi: 10.1161/CIRCULATIONAHA.117.029652

22 Stringhini S, Carmeli C, Jokela M, et al. Socioeconomic status and the 25 × 25 risk factors as determinants of premature mortality: a multicohort study and meta-analysis of 1·7 million men and women. The Lancet. 2017;389:1229–37. doi: 10.1016/S0140-6736(16)32380-7

23 Xiao H, Ding K, Li X, et al. A digital-health multidomain lifestyle management framework and its associations with cardiometabolic health: a real-world observational study. BMC Med. 2026;24:285. doi: 10.1186/s12916-026-04830-y

24 Kameda S, Makimoto H, Fujiwara T, et al. Impact of annual health check-ups on improvement in hypertension and abnormalities of glucose and lipid metabolism. Hypertens Res. 2026;49:516–25. doi: 10.1038/s41440-025-02465-9

25 Bauvin P, Benani A, Lepoittevin M, et al. A Comprehensive Prospective Cohort in Preventive Medicine: Protocol and Profile of the First 1,000 Participants in a Health Screening Program. 2025.

26 Lai H, Tu Y, Liao C, et al. Joint assessment of abdominal obesity and non-traditional lipid parameters for primary prevention of cardiometabolic multimorbidity: insights from the China health and retirement longitudinal study 2011-2018. Cardiovasc Diabetol. 2025;24:109. doi: 10.1186/s12933-025-02667-y

27 Chung F, Subramanyam R, Liao P, et al. High STOP-Bang score indicates a high probability of obstructive sleep apnoea. British Journal of Anaesthesia. 2012;108:768–75. doi: 10.1093/bja/aes022

28 Johns MW. A New Method for Measuring Daytime Sleepiness: The Epworth Sleepiness Scale. Sleep. 1991;14:540–5. doi: 10.1093/sleep/14.6.540

29 Greenland S, Senn SJ, Rothman KJ, et al. Statistical tests, P values, confidence intervals, and power: a guide to misinterpretations. Eur J Epidemiol. 2016;31:337–50. doi: 10.1007/s10654-016-0149-3

30 Amrhein V, Greenland S, McShane B. Scientists rise up against statistical significance. Nature. 2019;567:305–7. doi: 10.1038/d41586-019-00857-9

31 Frederiksen H, Hjelmborg J, Mortensen J, et al. Age trajectories of grip strength: cross-sectional and longitudinal data among 8,342 Danes aged 46 to 102. Ann Epidemiol. 2006;16:554–62. doi: 10.1016/j.annepidem.2005.10.006

32 Bohannon RW. Test-Retest Reliability of Measurements of Hand-Grip Strength Obtained by Dynamometry from Older Adults: A Systematic Review of Research in the PubMed Database. J Frailty Aging. 2017;6:83–7. doi: 10.14283/jfa.2017.8

33 Bohannon RW. Minimal clinically important difference for grip strength: a systematic review. J Phys Ther Sci. 2019;31:75–8. doi: 10.1589/jpts.31.75

34 Villareal DT, Chode S, Parimi N, et al. Weight Loss, Exercise, or Both and Physical Function in Obese Older Adults. New England Journal of Medicine. 2011;364:1218–29. doi: 10.1056/NEJMoa1008234

35 Rahimi K, Bidel Z, Nazarzadeh M, et al. Pharmacological blood pressure lowering for primary and secondary prevention of cardiovascular disease across different levels of blood pressure: an individual participant-level data meta-analysis. The Lancet. 2021;397:1625–36. doi: 10.1016/S0140-6736(21)00590-0

36 Barnett AG, van der Pols JC, Dobson AJ. Regression to the mean: what it is and how to deal with it. Int J Epidemiol. 2005;34:215–20. doi: 10.1093/ije/dyh299

37 Chang AM, Halter JB. Aging and insulin secretion. Am J Physiol Endocrinol Metab. 2003;284:E7–12. doi: 10.1152/ajpendo.00366.2002

38 Regitz-Zagrosek V, Oertelt-Prigione S, Seeland U, et al. Sex and gender differences in myocardial hypertrophy and heart failure. Circ J. 2010;74:1265–73. doi: 10.1253/circj.cj-10-0196

39 Bauvin P, Benani A, Lepoittevin M, et al. Zoī cohort, a prospective cohort with comprehensive phenotyping for preventive medicine in France, first 1000 participants: cohort profile. BMJ Open. 2026;16:e111119. doi: 10.1136/bmjopen-2025-111119

40 McCambridge J, Witton J, Elbourne DR. Systematic review of the Hawthorne effect: new concepts are needed to study research participation effects. J Clin Epidemiol. 2014;67:267–77. doi: 10.1016/j.jclinepi.2013.08.015

41 Tabagisme□: usage, envie d’arrêter et tentatives d’arrêt. Baromètre de Santé publique France : résultats de l’édition 2024 | Santé publique France. 2025. https://www.santepubliquefrance.fr/tabac/rapportsynthese/tabagisme-usage-envie-darreter-et-tentatives-darret-barometre-de-sante-publique-france-resultats-de (accessed 11 September 2026)

