## Supplementary File for "Cardiovascular and metabolic trajectories in men engaged in a multidimensional preventive care program: a prospective observational study"

### I. Supplementary Methods

#### 1. Rationale for the male-only analysis

The present analysis is the first step of a sex-stratified analysis plan for the cohort. Men and women differ in body composition and fat distribution, in the hormonal regulation of lipid and glucose metabolism, and in the trajectories of lipid parameters across the life course; the SCORE2 algorithm is sex-specific; and in women the interpretation of body composition, lipid and neuroendocrine changes over one year requires accounting for menopausal status and for cyclic variation of some markers. Stratifying a mixed cohort would equally avoid confounding by sex; separate reports were preferred because the outcome definitions and the interpretation of several markers differ between sexes. The analysis of women is in preparation.

#### 2. Detailed program description and preventive recommendations

The Zoī preventive program is a structured multidisciplinary care pathway designed to evaluate and monitor health status across multiple physiological domains. Participants enter the program either on their own initiative, as self-paying individuals, or through a program offered by their employer. They are followed by a multidisciplinary medical team including physicians specialized in general and cardiovascular preventive medicine.

During the baseline assessment day (CU1), participants undergo a standardized battery of clinical, biological, imaging and behavioral evaluations over approximately four hours: self-administered questionnaires, a clinical examination, anthropometric measurements, body composition analysis by dual-energy X-ray absorptiometry (DEXA), handgrip strength, seated blood pressure and resting heart rate, fasting venous blood sampling and salivary sampling for cortisol and DHEA.

Approximately four weeks after CU1, each participant attends a follow-up medical consultation during which a physician reviews all results with the participant. Individualized preventive recommendations are delivered in a shared decision-making approach and cover lifestyle advice (nutrition, physical activity), dietary supplements tailored to the individual profile, pharmacological treatment when clinically indicated, and referral for additional evaluation or monitoring. Pharmacological interventions include statin therapy according to ESC/EAS LDL cholesterol targets adjusted for individual risk, antihypertensive agents and antiplatelet therapy as appropriate. Dietary supplements include berberine for metabolic abnormalities (elevated fasting glucose or HbA1c, unfavorable body composition), omega-3 fatty acids for elevated triglycerides, coenzyme Q10 in participants receiving statins, magnesium for documented deficiency, vitamin B9 and methylcobalamin for elevated homocysteine or low red blood cell folate, and pregnenolone for documented neuroendocrine imbalance.

At approximately three months, a nurse-led follow-up call recalls the recommendations and addresses barriers. Between evaluations, participants have access to a smartphone application through which they can consult their results and recommendations, receive reminders and report whether recommendations have been followed. Implementation of the recommendations is reviewed by the physician at the second evaluation (CU2) and recorded as a followed/not followed status, without dose, duration or objective verification; in the present cohort this status was documented for a minority of recommendations and was therefore not analyzed. Engagement metrics of the application were not available for this analysis. The annual reassessment (CU2) follows the same standardized protocol as CU1.

#### 3. Questionnaires

STOP-BANG. The STOP-BANG questionnaire screens for the risk of obstructive sleep apnea with eight yes/no items scored 0–8: Snoring, Tiredness, Observed apnea, blood Pressure (self-reported items), and Body mass index >35 kg/m², Age >50 years, Neck circumference >40 cm, male Gender (items derived from measurements or demographics). Between the two evaluations, the wording and response format of the four self-reported items were revised to align with the original instrument (for example, the blood-pressure item moved from a history of hypertension to a current diagnosis or treatment of high blood pressure). The items are scored identically at both visits and were considered clinically equivalent, so that scores are compared between visits. As a sensitivity analysis, the change in the self-reported sub-score (S+T+O, 0–3) and in the objective sub-score (B+A+N+G+P, 0–5) was analyzed separately (Table S4). STOP-BANG is a screening instrument; polysomnography is not part of the preventive assessment.

Epworth Sleepiness Scale. Eight items scored 0–3, total 0–24, identical at both visits.

Perceived stress. Perceived stress was assessed at CU1 with a multi-item program questionnaire summarized as a score from 0 to 1 ⟦nombre d'items et construction du score⟧. This questionnaire was replaced by a different instrument between the two evaluations, with no common item; the stress scores were therefore not compared between visits.

#### 4. Biological measurements

Participants attended in the morning in a fasting state, after a light dinner the evening before (typically rice, white meat and water) and nothing but water afterwards. All blood and salivary analyses were performed by a single accredited clinical laboratory (Synlab) throughout the study period. The panel analyzed here comprised total, HDL and calculated LDL cholesterol, triglycerides, fasting glucose, insulin (pmol/L), HbA1c (NGSP, %), uric acid, homocysteine and high-sensitivity C-reactive protein; salivary cortisol at awakening, 30 minutes after awakening and in the evening, and evening DHEA.

Derived indices were recomputed from the constituent measurements of each participant: triglyceride/HDL ratio = triglycerides / HDL cholesterol; atherogenic index = total cholesterol / HDL cholesterol; non-HDL cholesterol = total − HDL cholesterol; HOMA-IR = fasting glucose (mg/dL) × fasting insulin (µU/mL) / 405, with insulin converted from pmol/L to µU/mL by division by 6.945. Cortisol awakening response = (cortisol at +30 min − cortisol at awakening) / cortisol at awakening × 100; awakening/evening ratio = cortisol at awakening / cortisol in the evening; cortisol/DHEA ratio = evening cortisol / evening DHEA.

SCORE2 was computed with the European Society of Cardiology algorithm for low-risk countries from age, sex, smoking status, systolic blood pressure and non-HDL cholesterol, at each evaluation with the age and measurements of that evaluation. SCORE2 is validated for ages 40–69 years; changes are reported for the whole cohort and for men aged 40–69 years at both evaluations.

HbA1c discontinuity. A step increase in HbA1c values of about 0.25 percentage points was observed in participants whose second evaluation took place from October 2025 onwards (Table S6), without a corresponding change in fasting glucose and with identical month-by-month medians whatever the data-recording pathway. The laboratory reported no change in method or calibration over the period, and the origin of the discontinuity could not be established. HbA1c results are therefore reported for the whole cohort and by period and are not interpreted as a biological change.

#### 5. Statistical analysis

Definitions. For each variable X, the within-individual change is ΔXᵢ = Xᵢ,CU2 − Xᵢ,CU1 in participants with a value at both evaluations (complete pairs; the number of pairs is given for every variable). Variables with fewer than 150 pairs were not analyzed longitudinally. The paired change is estimated by the Hodges–Lehmann estimator (the median of the Walsh averages (Δᵢ + Δⱼ)/2, i ≤ j) with its 95% confidence interval derived from the Wilcoxon signed-rank distribution, and tested with the two-sided Wilcoxon signed-rank test without continuity correction. The mean change with its t-based 95% CI and the paired Cohen's d (mean Δ / SD of Δ) are also reported. The relative change (Δ%) is the Hodges–Lehmann estimate divided by the CU1 median.

Plausibility ranges. Before analysis, values outside the following pre-specified ranges were set to missing: visceral adipose tissue mass 50–6 000 g; systolic blood pressure 70–250 mmHg; diastolic blood pressure 40–150 mmHg; resting heart rate 30–180 bpm; grip strength 5–90 kg; HDL cholesterol 10–150 mg/dL; LDL cholesterol 20–400 mg/dL; total cholesterol 80–500 mg/dL; triglycerides 20–1 500 mg/dL; fasting glucose 40–400 mg/dL; HbA1c 3.5–15%; body mass 35–200 kg; waist circumference 50–180 cm. Fourteen values from 11 participants were set to missing (four visceral adipose tissue values, one systolic and one diastolic blood pressure, two grip strength values, one HDL cholesterol and one triglyceride value, one body mass and three waist circumference values), all of them incompatible with life or with the measurement method and attributable to data-entry or unit errors.

Subgroup analyses (Table S3) describe the change within baseline categories of BMI, systolic blood pressure, glycemic status and LDL cholesterol with the same estimator and confidence interval, without hypothesis testing. Transitions (Table 3 and Table S8) use paired 2×2 tables: b is the number of participants moving from the unfavorable to the favorable category and c the reverse; the paired odds ratio is c/b with a Wald 95% CI on the log scale (Haldane–Anscombe correction of 0.5 when b or c is 0), the p-value is from the exact binomial McNemar test, and the net proportion improved is (b − c)/n.

Sensitivity analyses. Selection into follow-up: men with a second evaluation were compared with men whose first evaluation took place at least 15 months before data extraction but who had no second evaluation, using standardized mean differences (difference in means / pooled SD) and Wilcoxon rank-sum tests (Table S2). Plausible interval: the analyses of Table 2 were repeated in pairs whose second evaluation preceded the extraction date and whose interval was between 6 and 24 months (Table S5). Period: blood and salivary biomarkers were analyzed separately in men whose second evaluation took place before or from 1 October 2025 (Table S6). Interval: for each variable, the annualized change (Δ / interval in years, Hodges–Lehmann estimate) and the association between the individual change and the interval length (Spearman correlation; slope of a linear regression of Δ on the interval in years) are reported (Table S7).

Multiple testing. p-values were adjusted with the Benjamini–Hochberg false discovery rate procedure, applied globally over the 36 paired comparisons of Table 2 (families A to D together) and separately within Table 3 and within each supplementary table; raw p-values and adjusted q-values are shown side by side.

Sample size. No a priori sample size calculation was performed; the sample corresponds to all eligible men in the database at extraction. A posteriori, with a median of 232 pairs per variable, a two-sided paired test at α = 0.05 with 80% power detects a standardized within-individual difference d = (z₀.₉₇₅ + z₀.₈₀)/√n ≈ 0.18 (d ≈ 0.27 under a Bonferroni-type α of 0.05/36).

Software. All analyses were performed with R version ⟦x.y.z⟧ (R Foundation for Statistical Computing) using base R functions only; the analysis scripts and log are available from the corresponding author.

#### 6. Clinical categories, thresholds and formulas

Thresholds used in Table 3 are the fixed reference values in use at the center at the time of the study, applied identically to CU1 and CU2 values; laboratory interpretation flags, which can change between visits, were not used. Guideline-based thresholds (Table S8) are given as a sensitivity analysis.

| **Variable** | **Unfavorable if (Table 3)** | **Guideline-based threshold (Table S8)** |
| --- | --- | --- |
| Systolic blood pressure | ≥120 mmHg | ≥130 mmHg (ESC/ESH 2018 high-normal); ≥140 mmHg (hypertension) [1] |
| Android fat / tissue mass | ≥30.2% | — |
| Gynoid fat / tissue mass | ≥25.4% | — |
| Visceral adipose tissue mass | ≥800 g | same (internal consensus) |
| Waist circumference | ≥94 cm | — |
| Grip strength, right / left | ≤36.3 kg / ≤33.8 kg | — |
| HDL cholesterol | ≤46 mg/dL | ≤40 mg/dL (ESC/EAS 2019) [5] |
| Fasting glucose | ≥97 mg/dL | ≥100 mg/dL (ADA prediabetes) [3] |
| Total cholesterol | ≥191 mg/dL | — |
| LDL cholesterol | ≥115 mg/dL | ≥116 mg/dL (ESC/EAS 2019 target, low risk) [5] |
| Triglycerides | ≥114 mg/dL | ≥150 mg/dL (ESC/EAS 2019) [5] |
| Atherogenic index (TC/HDL) | ≥4.77 | — |
| Uric acid | ≥7.0 mg/dL | same (EULAR) [6] |
| HbA1c | ≥5.2% | ≥5.7% (ADA prediabetes) [3] |
| HOMA-IR | ≥1.93 | — |
| Insulin | ≥53 pmol/L | — |
| Cortisol awakening / evening ratio | ≤3.6 | — |
| DHEA, evening | ≤4.5 nmol/L | — |
| Cortisol, evening | ≥2.7 nmol/L | — |
| Cortisol / DHEA ratio, evening | ≥0.59 | — |
| Cortisol, +30 min | ≥11.0 nmol/L | — |
| Cortisol, awakening | ≥12.2 nmol/L | — |

Baseline categories used in Table S3: BMI <25, 25–29.9 and ≥30 kg/m² (WHO) [2]; systolic blood pressure <130, 130–139 and ≥140 mmHg (ESC/ESH 2018) [1]; glycemic status normoglycemic versus prediabetes/diabetes (fasting glucose ≥100 mg/dL or HbA1c ≥5.7%; ADA) [3]; LDL cholesterol <116, 116–189 and ≥190 mg/dL (ESC/EAS 2019) [5]. BMI = body mass / height²; HOMA-IR as in section 4 [4].

### II. Supplementary data

**Table S1. Variables analyzed longitudinally: units, method and number of paired observations (279 men)**

| **Variable (unit)** | **n pairs** |
| --- | --- |
| **A — Clinical and functional measures. Clinical examination; DEXA (body composition); digital dynamometer (grip strength); automated seated blood pressure** | |
| Body mass (kg) | 230 |
| Body fat mass (kg) | 231 |
| Body lean mass (kg) | 231 |
| Android fat / tissue mass (%) | 231 |
| Visceral adipose tissue mass (g) | 225 |
| Waist circumference (cm) | 196 |
| BMI (kg/m²) | 225 |
| Grip strength — right (kg) | 233 |
| Grip strength — left (kg) | 233 |
| Systolic BP (mmHg) | 245 |
| Diastolic BP (mmHg) | 245 |
| Heart rate, resting (bpm) | 247 |
| **B — Cardiometabolic blood biomarkers and SCORE2. Fasting venous blood, single clinical laboratory (Synlab); derived indices recomputed from constituents; SCORE2 computed from age, smoking, systolic BP and non-HDL cholesterol** | |
| Triglycerides (mg/dL) | 236 |
| HDL cholesterol (mg/dL) | 236 |
| LDL cholesterol (mg/dL) | 238 |
| Total cholesterol (mg/dL) | 238 |
| Non-HDL cholesterol (mg/dL) | 236 |
| Triglyceride / HDL ratio | 236 |
| Atherogenic index (total/HDL) | 236 |
| Fasting glucose (mg/dL) | 237 |
| Insulin (pmol/L) | 238 |
| HbA1c (%) | 231 |
| HOMA-IR | 237 |
| Homocysteine (µmol/L) | 233 |
| Uric acid (mg/dL) | 237 |
| hs-CRP (mg/L) | 155 |
| SCORE2, 10-year risk (%) | 250 |
| **C — Questionnaire scores. Self-administered questionnaires** | |
| STOP-BANG score (0–8) | 193 |
| Epworth sleepiness score (0–24) | 228 |
| **D — Neuroendocrine markers (saliva). Salivary samples at awakening, +30 min and evening** | |
| Cortisol, awakening (nmol/L) | 225 |
| Cortisol, +30 min (nmol/L) | 223 |
| Cortisol, evening (nmol/L) | 227 |
| Cortisol awakening response (%) | 221 |
| Cortisol awakening / evening ratio | 223 |
| DHEA, evening (nmol/L) | 223 |
| Cortisol / DHEA ratio, evening | 222 |

*n pairs = participants with a value at both evaluations after application of the plausibility ranges (Supplementary Methods, section 5). Derived indices (triglyceride/HDL, total/HDL, non-HDL, HOMA-IR) were recomputed from their constituents. Variables of the program not listed here were not analyzed longitudinally (fewer than 150 pairs or not part of the cardiometabolic objective).*

**Table S2. Baseline (CU1) characteristics of eligible men with and without a second evaluation**

| **Variable** | **n (with / without CU2)** | **Median, with CU2** | **Median, without CU2** | **Mean, with CU2** | **Mean, without CU2** | **SMD** | **p** |
| --- | --- | --- | --- | --- | --- | --- | --- |
| Age (years) | 279 / 584 | 51.2 | 49.1 | 50.0 | 49.7 | 0.026 | 0.41 |
| BMI (kg/m²) | 276 / 578 | 24.5 | 24.9 | 24.9 | 25.4 | −0.135 | 0.22 |
| Systolic BP (mmHg) | 279 / 567 | 127 | 127 | 127 | 128 | −0.052 | 0.73 |
| LDL cholesterol (mg/dL) | 279 / 569 | 126 | 126 | 127 | 126 | 0.018 | 0.77 |
| HDL cholesterol (mg/dL) | 279 / 571 | 56 | 55 | 57 | 57 | −0.059 | 0.89 |
| Fasting glucose (mg/dL) | 278 / 567 | 87 | 86 | 88 | 88 | 0.029 | 0.18 |
| HbA1c (%) | 279 / 571 | 5.20 | 5.20 | 5.26 | 5.26 | −0.003 | 0.99 |
| Visceral adipose tissue mass (g) | 260 / 543 | 996 | 1069 | 1251 | 1556 | −0.102 | 0.12 |
| Grip strength, right (kg) | 264 / 553 | 34.1 | 34.4 | 34.1 | 34.5 | −0.045 | 0.49 |
| SCORE2 (%) | 274 / 558 | 3.04 | 2.85 | 3.53 | 3.93 | −0.127 | 0.95 |
| Uric acid (mg/dL) | 279 / 571 | 6.10 | 6.20 | 6.09 | 6.23 | −0.135 | 0.16 |
| hs-CRP (mg/L) | 202 / 492 | 0.64 | 0.76 | 1.41 | 1.56 | −0.059 | 0.04 |

*Eligible men = first evaluation at least 15 months before extraction (n = 865: 279 with and 586 without a second evaluation). SMD = standardized mean difference (with − without) / pooled SD; p = Wilcoxon rank-sum test (unadjusted). All |SMD| ≤ 0.14.*

**Table S3. Within-individual changes according to baseline BMI, systolic blood pressure, glycemic status and LDL cholesterol categories**

| **Variable** | **Baseline category** | **n pairs** | **CU1, median [IQR]** | **Δ (Δ%)** | **95% CI** | **Cohen's d** |
| --- | --- | --- | --- | --- | --- | --- |
| **A — Body mass index and systolic blood pressure** | | | | | | |
| BMI (kg/m²) | BMI <25 kg/m² | 131 | 23.3 [21.8, 24.2] | 0.16 (0.7%) | [−0.03, 0.40] | 0.16 |
| BMI (kg/m²) | BMI 25–29.9 kg/m² | 79 | 26.3 [25.6, 27.5] | −0.14 (−0.5%) | [−0.55, 0.23] | −0.10 |
| BMI (kg/m²) | BMI ≥30 kg/m² | 15 | 32.0 [31.0, 34.3] | −1.75 (−5.5%) | [−2.69, −0.69] | −0.78 |
| Systolic BP (mmHg) | SBP <130 mmHg | 150 | 120 [113, 125] | 1.0 (0.8%) | [0.0, 2.5] | 0.15 |
| Systolic BP (mmHg) | SBP 130–139 mmHg | 61 | 135 [133, 137] | −3.5 (−2.6%) | [−5.5, −0.5] | −0.32 |
| Systolic BP (mmHg) | SBP ≥140 mmHg | 34 | 145 [141, 152] | −4.5 (−3.1%) | [−9.0, −1.0] | −0.46 |
| **B — Glycemic status and LDL cholesterol** | | | | | | |
| Fasting glucose (mg/dL) | Normoglycemic | 207 | 86 [81, 91] | −1.0 (−1.2%) | [−2.0, 0.0] | −0.12 |
| Fasting glucose (mg/dL) | Glucose ≥100 mg/dL or HbA1c ≥5.7% | 30 | 102 [95, 107] | −2.5 (−2.4%) | [−6.5, 0.5] | −0.18 |
| HbA1c (%) | Normoglycemic | 201 | 5.2 [5.0, 5.4] | 0.10 (1.9%) | [0.05, 0.15] | 0.32 |
| HbA1c (%) | Glucose ≥100 mg/dL or HbA1c ≥5.7% | 30 | 5.8 [5.3, 6.0] | 0.10 (1.7%) | [−0.05, 0.20] | 0.20 |
| LDL cholesterol (mg/dL) | LDL <116 mg/dL | 95 | 100 [88, 108] | 6.5 (6.5%) | [2.0, 11.0] | 0.32 |
| LDL cholesterol (mg/dL) | LDL 116–189 mg/dL | 133 | 138 [128, 158] | −2.0 (−1.4%) | [−7.5, 2.5] | −0.18 |
| LDL cholesterol (mg/dL) | LDL ≥190 mg/dL | 10 | 197 [195, 214] | −38.5 (−19.5%) | [−78.0, −4.0] | −0.83 |

*Descriptive analyses within baseline categories; Δ = Hodges–Lehmann estimate with 95% CI from the Wilcoxon signed-rank distribution; no hypothesis test. The LDL ≥190 mg/dL category rests on 10 observations, none of whom had a statin prescription recorded at CU1. HbA1c changes are affected by the period discontinuity described in Supplementary Methods, section 4, and Table S6.*

**Table S4. STOP-BANG: within-individual change of the total score and of the self-reported and objective sub-scores**

| **Score** | **n pairs** | **CU1, median [IQR]** | **CU1, mean (SD)** | **CU2, median** | **Δ, Hodges–Lehmann [95% CI]** | **Δ, mean [95% CI]** | **Cohen's d** | **p** | **n with change** |
| --- | --- | --- | --- | --- | --- | --- | --- | --- | --- |
| STOP-BANG total (0-8) | 193 | 3 [2, 4] | 2.95 (1.15) | 2 | −1.0 [−1.0, −1.0] | −0.59 [−0.73, −0.45] | −0.59 | <0.001 | 123 |
| Self-reported items S+T+O (0-3) | 193 | 1 [1, 2] | 1.20 (0.84) | 0 | −1.0 [−1.0, −1.0] | −0.55 [−0.68, −0.43] | −0.63 | <0.001 | 121 |
| Objective items B+A+N+G+P (0-5) | 193 | 2 [1, 2] | 1.75 (0.73) | 2 | 0.0 [−1.0, 0.0] | −0.04 [−0.10, 0.02] | −0.09 | 0.24 | 32 |

*S+T+O = snoring, tiredness, observed apnea (self-reported items whose wording was revised between the visits); B+A+N+G+P = BMI >35 kg/m², age >50 years, neck circumference >40 cm, male gender, blood pressure. The decrease in the total score is entirely carried by the self-reported items; the Hodges–Lehmann CI of the total and S+T+O scores is degenerate because almost all non-zero individual changes equal −1. p = Wilcoxon signed-rank test (not adjusted).*

**Table S5. Sensitivity analysis: within-individual changes in pairs with a plausible interval between evaluations (6–24 months, CU2 before the extraction date)**

| **Variable** | **n pairs** | **Δ (Δ%)** | **95% CI** | **Cohen's d** | **p** | **q** |
| --- | --- | --- | --- | --- | --- | --- |
| **A — Clinical and functional measures** | | | | | | |
| Body mass (kg) | 226 | −0.55 (−0.7%) | [−0.95, −0.10] | −0.20 | 0.01 | 0.05 |
| Body fat mass (kg) | 227 | −0.40 (−1.9%) | [−0.74, −0.08] | −0.16 | 0.01 | 0.05 |
| Body lean mass (kg) | 227 | 0.08 (0.1%) | [−0.14, 0.29] | 0.08 | 0.45 | 0.54 |
| Android fat / tissue mass (%) | 227 | −0.55 (−1.7%) | [−1.05, −0.05] | −0.18 | 0.03 | 0.08 |
| Visceral adipose tissue mass (g) | 221 | −49 (−5.0%) | [−80, −18] | −0.24 | 0.002 | 0.007 |
| Waist circumference (cm) | 192 | 0.0 (0.0%) | [−1.0, 0.5] | −0.05 | 0.95 | 0.95 |
| BMI (kg/m²) | 217 | −0.02 (−0.1%) | [−0.28, 0.17] | −0.04 | 0.70 | 0.75 |
| Grip strength — right (kg) | 228 | 2.40 (7.0%) | [1.70, 3.00] | 0.48 | <0.001 | <0.001 |
| Grip strength — left (kg) | 228 | 1.70 (5.2%) | [1.05, 2.30] | 0.23 | <0.001 | <0.001 |
| Systolic BP (mmHg) | 239 | −0.5 (−0.4%) | [−1.5, 0.5] | −0.06 | 0.41 | 0.53 |
| Diastolic BP (mmHg) | 239 | 0.0 (0.0%) | [−1.0, 0.5] | −0.03 | 0.76 | 0.79 |
| Heart rate, resting (bpm) | 241 | −0.5 (−0.8%) | [−1.5, 0.5] | −0.09 | 0.19 | 0.31 |
| **B — Cardiometabolic blood biomarkers and SCORE2** | | | | | | |
| Triglycerides (mg/dL) | 231 | −3.0 (−3.3%) | [−7.0, 1.0] | −0.13 | 0.13 | 0.23 |
| HDL cholesterol (mg/dL) | 231 | 3.5 (6.3%) | [2.5, 4.5] | 0.43 | <0.001 | <0.001 |
| LDL cholesterol (mg/dL) | 233 | 1.0 (0.8%) | [−2.5, 4.5] | −0.05 | 0.55 | 0.63 |
| Total cholesterol (mg/dL) | 233 | 3.5 (1.7%) | [−0.5, 7.5] | 0.03 | 0.08 | 0.20 |
| Non-HDL cholesterol (mg/dL) | 233 | 0.5 (0.3%) | [−3.0, 4.0] | −0.07 | 0.70 | 0.75 |
| Triglyceride / HDL ratio | 232 | −0.15 (−9.6%) | [−0.24, −0.07] | −0.26 | <0.001 | 0.002 |
| Atherogenic index (total/HDL) | 233 | −0.13 (−3.7%) | [−0.22, −0.06] | −0.28 | <0.001 | 0.002 |
| Fasting glucose (mg/dL) | 232 | −1.0 (−1.1%) | [−2.0, 0.0] | −0.13 | 0.02 | 0.06 |
| Insulin (pmol/L) | 233 | −1.0 (−3.1%) | [−3.0, 1.0] | −0.05 | 0.33 | 0.46 |
| HbA1c (%) | 226 | 0.10 (1.9%) | [0.05, 0.15] | 0.22 | <0.001 | <0.001 |
| HOMA-IR | 231 | −0.04 (−4.1%) | [−0.10, 0.02] | −0.07 | 0.18 | 0.30 |
| Homocysteine (µmol/L) | 228 | 0.0 (0.0%) | [0.0, 0.5] | 0.04 | 0.43 | 0.54 |
| Uric acid (mg/dL) | 232 | −0.10 (−1.6%) | [−0.20, 0.00] | −0.10 | 0.11 | 0.21 |
| hs-CRP (mg/L) | 152 | −0.13 (−19.4%) | [−0.26, −0.03] | −0.20 | 0.01 | 0.05 |
| SCORE2, 10-year risk (%) | 238 | −0.06 (−1.9%) | [−0.12, 0.01] | −0.10 | 0.10 | 0.20 |
| **D — Neuroendocrine markers (saliva)** | | | | | | |
| Cortisol, awakening (nmol/L) | 220 | 0.25 (3.5%) | [−0.45, 0.95] | 0.02 | 0.49 | 0.58 |
| Cortisol, +30 min (nmol/L) | 218 | −0.40 (−3.7%) | [−1.25, 0.45] | −0.05 | 0.34 | 0.46 |
| Cortisol, evening (nmol/L) | 222 | −0.25 (−27.8%) | [−0.55, 0.05] | −0.05 | 0.10 | 0.20 |
| Cortisol awakening response (%) | 216 | −8.0 (−20.8%) | [−22.5, 6.0] | 0.06 | 0.25 | 0.37 |
| Cortisol awakening / evening ratio | 218 | 7.5 (89.3%) | [2.0, 15.6] | 0.18 | 0.002 | 0.007 |
| DHEA, evening (nmol/L) | 218 | −0.45 (−5.6%) | [−1.20, 0.25] | −0.07 | 0.21 | 0.32 |
| Cortisol / DHEA ratio, evening | 217 | −0.025 (−27.8%) | [−0.060, 0.005] | −0.05 | 0.10 | 0.20 |

*Pairs excluded: 27 second evaluations dated after the extraction date, 2 intervals <6 months and 11 intervals >24 months (n = 239 men). Δ = Hodges–Lehmann estimate; q = Benjamini–Hochberg adjusted p-value over the tests of this table. Questionnaire scores were not recomputed in this subset.*

**Table S6. Blood and salivary biomarkers: within-individual changes by period of the second evaluation (before versus from 1 October 2025)**

| **Variable** | **n pairs, before Oct 2025** | **Δ [95% CI], before Oct 2025** | **p** | **n pairs, from Oct 2025** | **Δ [95% CI], from Oct 2025** | **p** |
| --- | --- | --- | --- | --- | --- | --- |
| Systolic BP (mmHg) | 125 | −1.5 [−3.0, 0.0] | 0.07 | 120 | 0.5 [−1.5, 2.0] | 0.74 |
| Diastolic BP (mmHg) | 125 | −1.0 [−2.5, 0.0] | 0.10 | 120 | 0.5 [−0.5, 2.0] | 0.35 |
| Triglycerides (mg/dL) | 123 | −3.0 [−9.0, 3.0] | 0.32 | 113 | −3.5 [−9.0, 2.0] | 0.21 |
| HDL cholesterol (mg/dL) | 123 | 4.0 [3.0, 5.5] | <0.001 | 113 | 2.5 [1.0, 4.5] | <0.001 |
| LDL cholesterol (mg/dL) | 125 | 0.0 [−5.5, 5.0] | 0.99 | 113 | 1.5 [−3.0, 6.0] | 0.49 |
| Total cholesterol (mg/dL) | 125 | 3.0 [−3.0, 9.0] | 0.32 | 113 | 3.0 [−2.0, 8.5] | 0.21 |
| Non-HDL cholesterol (mg/dL) | 125 | 0.0 [−5.5, 5.0] | 0.99 | 113 | 1.0 [−4.0, 5.5] | 0.73 |
| Triglyceride / HDL ratio | 125 | −0.17 [−0.31, −0.04] | 0.007 | 112 | −0.15 [−0.26, −0.04] | 0.009 |
| Atherogenic index (total/HDL) | 125 | −0.18 [−0.30, −0.07] | <0.001 | 113 | −0.10 [−0.21, 0.01] | 0.07 |
| Fasting glucose (mg/dL) | 124 | −2.0 [−3.0, −0.5] | 0.004 | 113 | 0.0 [−1.5, 1.0] | 0.88 |
| Insulin (pmol/L) | 125 | −2.5 [−5.0, 0.0] | 0.06 | 113 | 1.0 [−2.0, 3.5] | 0.55 |
| HbA1c (%) | 125 | −0.05 [−0.15, 0.00] | 0.003 | 106 | 0.25 [0.25, 0.30] | <0.001 |
| HOMA-IR | 124 | −0.09 [−0.18, −0.01] | 0.02 | 112 | 0.02 [−0.07, 0.12] | 0.70 |
| Homocysteine (µmol/L) | 120 | 0.5 [0.0, 1.5] | 0.04 | 113 | 0.0 [−1.0, 0.5] | 0.44 |
| Uric acid (mg/dL) | 124 | −0.10 [−0.25, 0.00] | 0.10 | 113 | −0.05 [−0.20, 0.10] | 0.43 |
| hs-CRP (mg/L) | 58 | −0.08 [−0.22, 0.07] | 0.22 | 97 | −0.17 [−0.44, −0.01] | 0.03 |
| Cortisol, awakening (nmol/L) | 122 | 0.80 [0.00, 1.70] | 0.06 | 103 | −0.40 [−1.60, 0.70] | 0.47 |
| Cortisol, +30 min (nmol/L) | 121 | 0.30 [−0.80, 1.40] | 0.58 | 102 | −1.25 [−2.45, 0.20] | 0.08 |
| Cortisol, evening (nmol/L) | 121 | −0.30 [−0.65, 0.10] | 0.19 | 106 | −0.30 [−0.70, 0.20] | 0.26 |
| Cortisol awakening response (%) | 120 | −14.5 [−34.1, 5.1] | 0.14 | 101 | −1.1 [−23.9, 21.1] | 0.93 |
| Cortisol awakening / evening ratio | 120 | 6.2 [0.6, 17.7] | 0.03 | 103 | 8.3 [1.1, 20.0] | 0.02 |
| DHEA, evening (nmol/L) | 122 | −0.40 [−1.40, 0.65] | 0.45 | 101 | −0.45 [−1.60, 0.60] | 0.38 |
| Cortisol / DHEA ratio, evening | 121 | −0.035 [−0.085, 0.010] | 0.12 | 101 | −0.025 [−0.075, 0.025] | 0.29 |

*Δ = Hodges–Lehmann estimate with 95% CI; p = Wilcoxon signed-rank test (unadjusted). The HbA1c increase is confined to the later period (+0.25 [0.25, 0.30]) and absent before (−0.05 [−0.15, 0.00]); in men evaluated before October 2025 the proportion with HbA1c ≥5.2% was 55 (44.0%) at CU1 and 50 (40.0%) at CU2 (b = 12, c = 7, OR 0.58 [0.23, 1.48]). Systolic and diastolic blood pressure are shown for comparison. Changes in fasting glucose, insulin and HOMA-IR are also confined to the earlier period; these period differences are described without interpretation.*

**Table S7. Influence of the interval between evaluations: annualized changes and association between individual change and interval length**

| **Variable** | **n** | **Annualized Δ [95% CI] (per year)** | **Spearman ρ (Δ vs interval)** | **p** | **β (Δ per year of interval)** | **p** |
| --- | --- | --- | --- | --- | --- | --- |
| **A — Clinical and functional measures** | | | | | | |
| Body mass (kg) | 230 | −0.48 [−0.87, −0.10] | 0.02 | 0.80 | −0.01 | 0.99 |
| Body fat mass (kg) | 231 | −0.36 [−0.66, −0.08] | −0.01 | 0.88 | −0.45 | 0.56 |
| Body lean mass (kg) | 231 | 0.07 [−0.12, 0.25] | 0.01 | 0.85 | −0.82 | 0.37 |
| Android fat / tissue mass (%) | 231 | −0.50 [−0.94, −0.06] | −0.03 | 0.70 | −0.76 | 0.49 |
| Visceral adipose tissue mass (g) | 225 | −43 [−69, −17] | −0.01 | 0.84 | −15 | 0.83 |
| Waist circumference (cm) | 196 | 0.0 [−0.7, 0.6] | 0.00 | 1.00 | 1.9 | 0.16 |
| BMI (kg/m²) | 224 | −0.02 [−0.20, 0.15] | 0.07 | 0.29 | 0.31 | 0.17 |
| Grip strength — right (kg) | 233 | 2.06 [1.46, 2.62] | 0.04 | 0.58 | −0.06 | 0.96 |
| Grip strength — left (kg) | 233 | 1.53 [0.95, 2.10] | −0.08 | 0.20 | −3.25 | 0.03 |
| Systolic BP (mmHg) | 245 | −0.6 [−1.6, 0.5] | 0.00 | 0.94 | 2.5 | 0.20 |
| Diastolic BP (mmHg) | 245 | −0.1 [−0.9, 0.6] | −0.06 | 0.36 | 0.7 | 0.64 |
| Heart rate, resting (bpm) | 247 | −0.6 [−1.4, 0.2] | 0.02 | 0.74 | 0.3 | 0.81 |
| **B — Cardiometabolic blood biomarkers and SCORE2** | | | | | | |
| Triglycerides (mg/dL) | 236 | −3.1 [−6.8, 0.4] | 0.10 | 0.14 | 5.5 | 0.37 |
| HDL cholesterol (mg/dL) | 236 | 3.0 [2.1, 4.0] | 0.13 | 0.05 | 3.6 | 0.04 |
| LDL cholesterol (mg/dL) | 238 | 0.6 [−2.5, 3.5] | 0.09 | 0.17 | 9.8 | 0.13 |
| Total cholesterol (mg/dL) | 238 | 2.4 [−1.0, 5.8] | 0.13 | 0.05 | 14.3 | 0.05 |
| Non-HDL cholesterol (mg/dL) | 238 | 0.1 [−3.1, 3.1] | 0.10 | 0.14 | 10.9 | 0.11 |
| Triglyceride / HDL ratio | 237 | −0.14 [−0.21, −0.07] | 0.02 | 0.70 | 0.02 | 0.87 |
| Atherogenic index (total/HDL) | 238 | −0.12 [−0.19, −0.06] | 0.01 | 0.94 | 0.06 | 0.70 |
| Fasting glucose (mg/dL) | 237 | −0.9 [−1.7, −0.1] | −0.10 | 0.13 | −1.8 | 0.29 |
| Insulin (pmol/L) | 238 | −0.6 [−2.2, 1.1] | −0.09 | 0.17 | −5.1 | 0.16 |
| HbA1c (%) | 231 | 0.09 [0.05, 0.13] | −0.02 | 0.80 | 0.09 | 0.25 |
| HOMA-IR | 236 | −0.03 [−0.08, 0.02] | −0.11 | 0.09 | −0.21 | 0.13 |
| Homocysteine (µmol/L) | 233 | 0.0 [−0.3, 0.4] | 0.17 | 0.01 | 1.7 | 0.003 |
| Uric acid (mg/dL) | 237 | −0.08 [−0.17, 0.00] | −0.05 | 0.47 | −0.16 | 0.29 |
| hs-CRP (mg/L) | 155 | −0.11 [−0.22, −0.02] | −0.07 | 0.41 | −0.10 | 0.75 |
| SCORE2, 10-year risk (%) | 250 | −0.04 [−0.10, 0.01] | 0.00 | 0.96 | 0.02 | 0.90 |
| **D — Neuroendocrine markers (saliva)** | | | | | | |
| Cortisol, awakening (nmol/L) | 225 | 0.30 [−0.30, 0.89] | −0.09 | 0.19 | −1.48 | 0.10 |
| Cortisol, +30 min (nmol/L) | 223 | −0.34 [−1.08, 0.40] | 0.00 | 0.96 | 1.73 | 0.14 |
| Cortisol, evening (nmol/L) | 227 | −0.27 [−0.51, 0.01] | 0.02 | 0.81 | −0.58 | 0.20 |
| Cortisol awakening response (%) | 221 | −8.1 [−21.0, 4.5] | 0.04 | 0.53 | −107.0 | 0.75 |
| Cortisol awakening / evening ratio | 223 | 6.7 [2.0, 13.4] | −0.09 | 0.19 | −5.7 | 0.55 |
| DHEA, evening (nmol/L) | 223 | −0.39 [−1.04, 0.25] | −0.01 | 0.87 | −0.91 | 0.57 |
| Cortisol / DHEA ratio, evening | 222 | −0.027 [−0.058, 0.001] | 0.04 | 0.56 | −0.040 | 0.64 |

*Annualized Δ = Hodges–Lehmann estimate of Δ / interval (years); ρ = Spearman correlation between the individual change and the interval length; β = slope of the linear regression of Δ on the interval (units of the variable per year). p-values unadjusted. Changes were unrelated to the interval for almost all variables (exceptions: homocysteine, HDL and total cholesterol, left-hand grip strength).*

**Table S8. Transitions across guideline-based categories between CU1 and CU2 with paired odds ratios**

| **Variable** | **Unfavorable if** | **Source of threshold** | **n** | **Unfavorable at CU1, n (%)** | **Unfavorable at CU2, n (%)** | **b / c** | **OR (c/b) [95% CI]** | **p** | **q** |
| --- | --- | --- | --- | --- | --- | --- | --- | --- | --- |
| Systolic BP (mmHg) | ≥ 130 | ESC/ESH 2018 high-normal | 245 | 95 (38.8%) | 86 (35.1%) | 28 / 19 | 0.68 [0.38, 1.22] | 0.24 | 0.39 |
| Systolic BP (mmHg) | ≥ 140 | ESC/ESH 2018 hypertension | 245 | 34 (13.9%) | 36 (14.7%) | 14 / 16 | 1.14 [0.56, 2.34] | 0.86 | 0.86 |
| Visceral adipose tissue mass (g) | ≥ 800 | internal consensus (as submitted) | 225 | 128 (56.9%) | 122 (54.2%) | 13 / 7 | 0.54 [0.21, 1.35] | 0.26 | 0.39 |
| HDL cholesterol (mg/dL) | ≤ 40 | ESC/EAS 2019 low HDL (<40 men) | 236 | 20 (8.5%) | 12 (5.1%) | 11 / 3 | 0.27 [0.08, 0.98] | 0.06 | 0.26 |
| Fasting glucose (mg/dL) | ≥ 100 | ADA prediabetes | 237 | 22 (9.3%) | 16 (6.8%) | 11 / 5 | 0.45 [0.16, 1.31] | 0.21 | 0.39 |
| LDL cholesterol (mg/dL) | ≥ 116 | ESC/EAS 2019 target low risk (<116) | 238 | 143 (60.1%) | 138 (58.0%) | 31 / 26 | 0.84 [0.50, 1.41] | 0.60 | 0.67 |
| Triglycerides (mg/dL) | ≥ 150 | ESC/EAS 2019 hypertriglyceridemia | 236 | 28 (11.9%) | 22 (9.3%) | 16 / 10 | 0.62 [0.28, 1.38] | 0.33 | 0.42 |
| Uric acid (mg/dL) | ≥ 7 | EULAR hyperuricemia | 237 | 56 (23.6%) | 42 (17.7%) | 30 / 16 | 0.53 [0.29, 0.98] | 0.05 | 0.26 |
| HbA1c (%) | ≥ 5.7 | ADA prediabetes | 231 | 15 (6.5%) | 22 (9.5%) | 4 / 11 | 2.75 [0.88, 8.64] | 0.12 | 0.36 |

*b = unfavorable at CU1 → favorable at CU2; c = the reverse; OR = c/b with Wald 95% CI; p = exact McNemar test; q = Benjamini–Hochberg adjusted within the table. Sources: ESC/ESH 2018 [1], WHO [2], ADA [3], ESC/EAS 2019 [5], EULAR [6]. The HbA1c transition is affected by the period discontinuity (Supplementary Methods, section 4).*

### Supplementary references

1. Williams B, Mancia G, Spiering W, et al. 2018 ESC/ESH Guidelines for the management of arterial hypertension. Eur Heart J. 2018;39(33):3021-3104.

2. World Health Organization. Obesity: preventing and managing the global epidemic. Report of a WHO Consultation. WHO Technical Report Series No. 894. Geneva: WHO; 2000.

3. American Diabetes Association Professional Practice Committee. Standards of Care in Diabetes—2024. Diabetes Care. 2024;47(Suppl 1):S1-S321.

4. Matthews DR, Hosker JP, Rudenski AS, Naylor BA, Treacher DF, Turner RC. Homeostasis model assessment: insulin resistance and beta-cell function from fasting plasma glucose and insulin concentrations in man. Diabetologia. 1985;28(7):412-419.

5. Mach F, Baigent C, Catapano AL, et al. 2019 ESC/EAS Guidelines for the management of dyslipidaemias: lipid modification to reduce cardiovascular risk. Eur Heart J. 2020;41(1):111-188.

6. Richette P, Doherty M, Pascual E, et al. 2016 updated EULAR evidence-based recommendations for the management of gout. Ann Rheum Dis. 2017;76(1):29-42.

7. SCORE2 working group and ESC Cardiovascular risk collaboration. SCORE2 risk prediction algorithms: new models to estimate 10-year risk of cardiovascular disease in Europe. Eur Heart J. 2021;42(25):2439-2454.

8. Chung F, Yegneswaran B, Liao P, et al. STOP questionnaire: a tool to screen patients for obstructive sleep apnea. Anesthesiology. 2008;108(5):812-821.

9. Johns MW. A new method for measuring daytime sleepiness: the Epworth sleepiness scale. Sleep. 1991;14(6):540-545.
